# Global Approaches to Older Abuse Research in Institutional Care Settings: A Systematic Review

**DOI:** 10.1101/2023.08.09.23293921

**Authors:** Maria Agaliotis, Tracey Morris, Ilan Katz, David Greenfield

**Author notes:** ^¶^All authors have made substantial contributions to all three of sections (1), (2) and (3) below. (1) the conception and design of the study, or acquisition of data, or analysis and interpretation of data: AGALIOTIS, MORRIS, KATZ, GREENFIELD. (2) drafting the article or revising it critically for important intellectual content: AGALIOTIS, MORRIS, KATZ, GREENFIELD. (3) final approval of the version to be submitted: AGALIOTIS, MORRIS, KATZ, GREENFIELD. Data Availability Statement: All relevant data are within the manuscript and its Supporting Information files. Ethical standards: This project did not collect empirical data so ethical approval was not required.

## Abstract

**Objective:** Over the last two decades, authors have argued the rate of abuse among older adults in institutional settings has been underestimated due to challenges in defining and responding to the issue. The purpose of this systematic review is to provide an in-depth analysis of empirical studies examining methodologies measuring abuse of older people residing in a long-term institutional care facility (nursing homes, independent living and assisted living facilities), specifically staff-to-resident abuse.

**Methods:** Guided by PRISMA guidelines, 10 databases were searched from 2005 till July 2023. This review inclusion criteria were any type of abuse, as defined by the World Health Organization reported by staff and residents, family and relatives, and public anonymous registries. This article also includes a methodological critical assessment of studies which has not been conducted before. To direct the review, we use four guiding questions: a) what are the study characteristics? b) what are the methods and measurement tools that have been used? c) what has been the impact of methodology on the results? and d) what is the quality of these studies?

**Results:** In the last 18 years, 22 studies from eight counties undertook cross-sectional examinations of staff-to-resident abuse. The review identified a heterogeneity of definitions of abuse and variations with who reported abuse, measurement tools and recall periods. We found the quality of studies varied significantly, with no consistency.

**Discussion:** These variations in study methodologies impacted the ability to synthesise the findings making it difficult to estimate a global prevalence rate of aged care abuse. From the analysis, we develop an Aged Care Abuse Research Checklist (ACARC) as a first step towards achieving a global standardized, evidence-based methodology for this field. Doing so will normalize processes within organizations and the community, allowing early interventions to change practices, reduce the risk of recurrence and improve resident quality of care and workplace cultures.

**Systematic Review Registration Number:** PROSPERO registry number: CRD42018055484, https://www.crd.york.ac.uk/PROSPERO.

## Introduction

Older people have higher risks of isolation, fragility, impaired cognitive function, and lack of social support structures; individually, and collectively, these issues make them vulnerable to maltreatment or abuse, most often from persons in trusting relationships [1]. Maltreatment and abuse can contribute to long term physical and psychological harms including stress, injury, depression, and increased mortality [2].

Recent Organization for Economic Cooperation and Development (OECD) data estimates between 6 – 20% of people aged 80 and over currently reside in institutional settings, and by 2050 this is likely to double [3]. This change is in part driven by the fact that the global population of people aged 60 and over, will increase from 962 million in 2017, to 2.1 billion by 2050 [4]. Institutional settings can range from independent living facilities, assisted living communities, nursing homes and continuing care retirement facilities. Abuse can be committed by staff-to-resident, resident-to-resident, or visitor-to-resident [5].

A 2019 systematic literature review found two in three residential unit staff self-reported committing abuse in the last year [5]. We know rates of abuse are reported to be higher among the vulnerable dependent older adults living in institutional settings, compared to older people in the general community [5], and yet many instances go unreported. Although evidence of extensive abuse of older adults is well established, challenges in defining, identifying, and responding to it restrict our ability to address the issue. In 2002, some clarity was brought to the problem by the World Health Organization (WHO), defining older adult abuse as ‘elder abuse’, and described it as an intentional or inappropriate act, single or repeated, causing distress or harm to an older adult [6]. Types of abuse include physical, psychological, or emotional, financial (or financial exploitation), sexual and neglect, intentional or unintentional [6]. Over the last ten years, there have been consistent calls to understand how to standardize, measure rates of abuse among older adults [7-10].

We know that over the last two decades, due to varying definitions and social norms across the world, the rate of abuse among older adults in institutional settings has been underestimated [10-13]. In short, our understanding of the prevalence of abuse of older adults is significantly limited and recent descriptions of instruments used to examine staff-to-resident abuse in residential care settings need a more thorough standardized investigation, since reporting abuse is as an essential part of public health, and reports of abuse is the responsibility of all members of the community [9, 10, 12]. Understanding the quality of abuse measurement tools among older adults [10, 12] within all aged care institutional settings, will provide a clearer picture of the how to better standardize the methodological approaches to measuring older age abuse in institutional settings.

This review in addition conducts a methodological quality assessment on empirical studies among all institutional settings, examining all modes of reporting older age abuse (staff, resident, relatives, or community [via registries including whether allegations or sustained acts of abuse]). Overall, the study aimed to investigate and develop a common standard research criteria to advance the methodological rigor in and practical viability approaches when measuring older abuse within institutional settings. Four guiding questions direct the review: (1) what are the study characteristics? (2) what are the methods and measurement tools that have been used and are they valid and reliable? (3) what has been the impact of methodology on the results? and (4) what is the level of quality of these studies?

## Methods

### Search strategy

A systematic quantitative review protocol was developed according to the PRISMA [14] (S1 Fig) and registered (SYSTEMATIC REVIEW REGISTRATION NUMBER: PROSPERO registry number: CRD42018055484, https://www.crd.york.ac.uk/PROSPERO)[15]. Ten academic databases (S1 File) were searched. The keyword search was informed by Lindbloom et al. (2007) [13] and a Cochrane review by Baker et al.(2016) [16] (S2 File). In addition to this search, full paper copies of potentially relevant articles were retrieved, and their reference lists screened.

### Eligibility criteria

The inclusion criteria included: observational studies reporting any incidence or prevalence data on any type of abuse as defined by the WHO (2022) (physical, psychological, financial, sexual abuse and neglect); as observed or committed abuse on older participants residing in long term institutional care facilities including assisted, independent or extended living facilities or care units, and residential or a nursing home; staff-to-resident abuse from ‘health care professional’ or ‘staff member’ to ‘patient’ or ‘resident’. Research articles were limited to full-text English language and published from 2005 till May 2020. This timing coincides with the last systematic review on abuse among older residents residing in nursing homes conducted by Lindbloom et al (2007). Additional searches were conducted using the same academic databases to retrieve studies published between May 2020 to July 2023. We also excluded studies based on study design such as single case reports; case series; and discussion or opinion pieces.

### Data Extraction and Data analysis

Titles and abstracts obtained from the search were screened by two reviewers (MA and TM). Duplicate articles were excluded. Data were extracted by one reviewer (TM) and independently audited by a second (MA). The data extraction was guided by an analytical framework using the elements of epidemiological methodology used in prevalence studies [17]. The framework characteristics and elements form the header columns for presented tables (Table 1) and rows form the information extracted from each article. Disagreement or ambiguities were resolved by consensus. Descriptive tables were developed based on the study recruitment methodology, that is who reported the abuse (staff, residents, relatives, or community) (S3 File). The subheading columns were structured based on the examining, study characteristics, methodology characteristics and results (S3 File).

**Table I:**
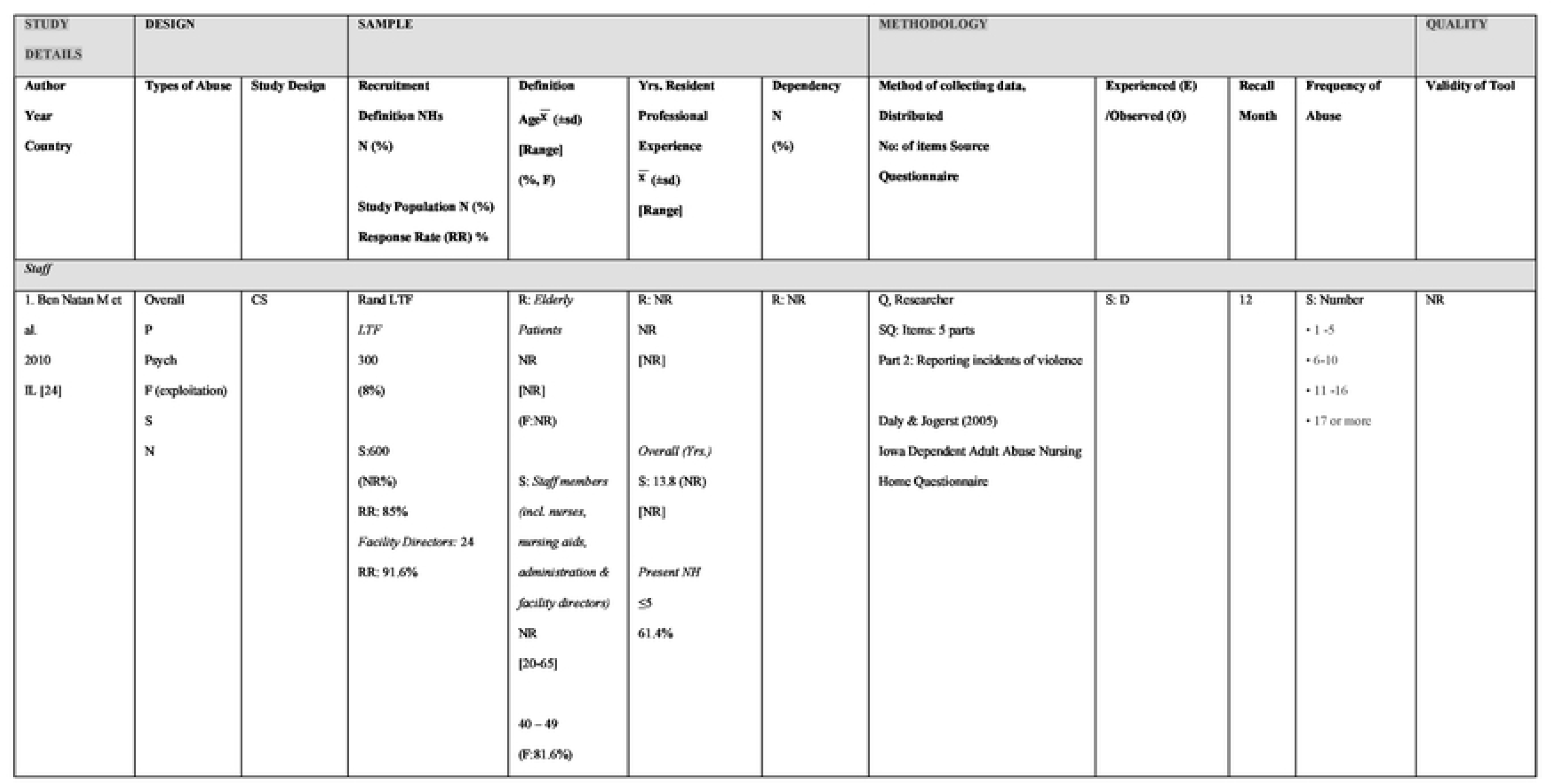
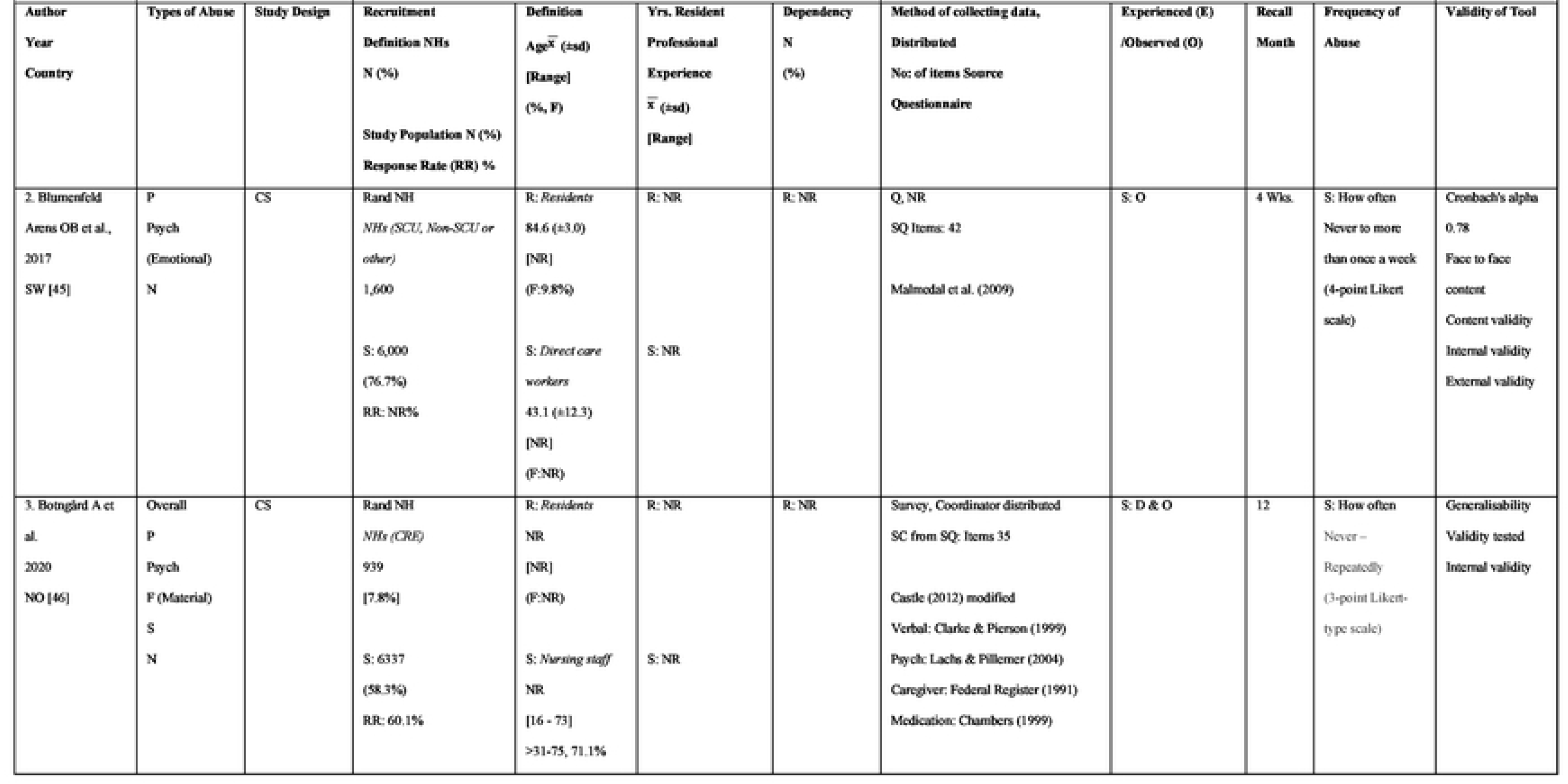
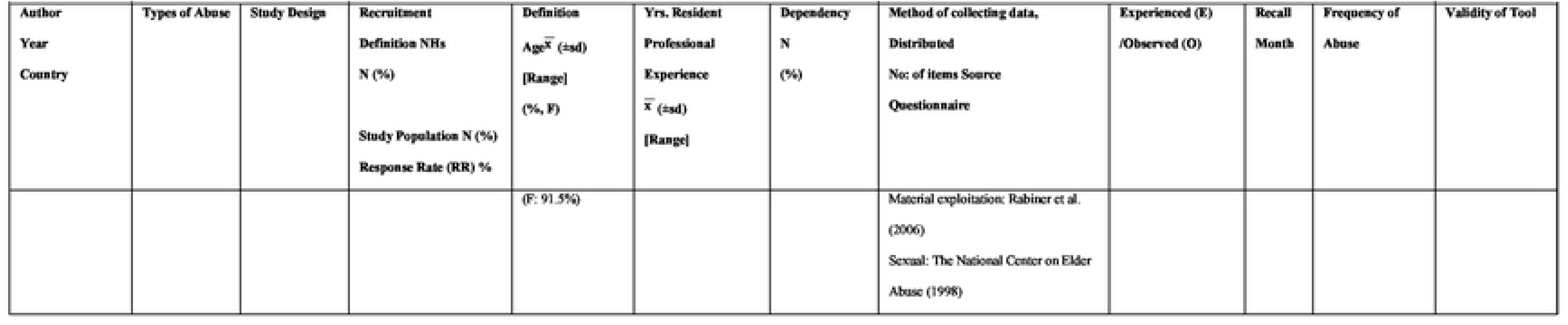
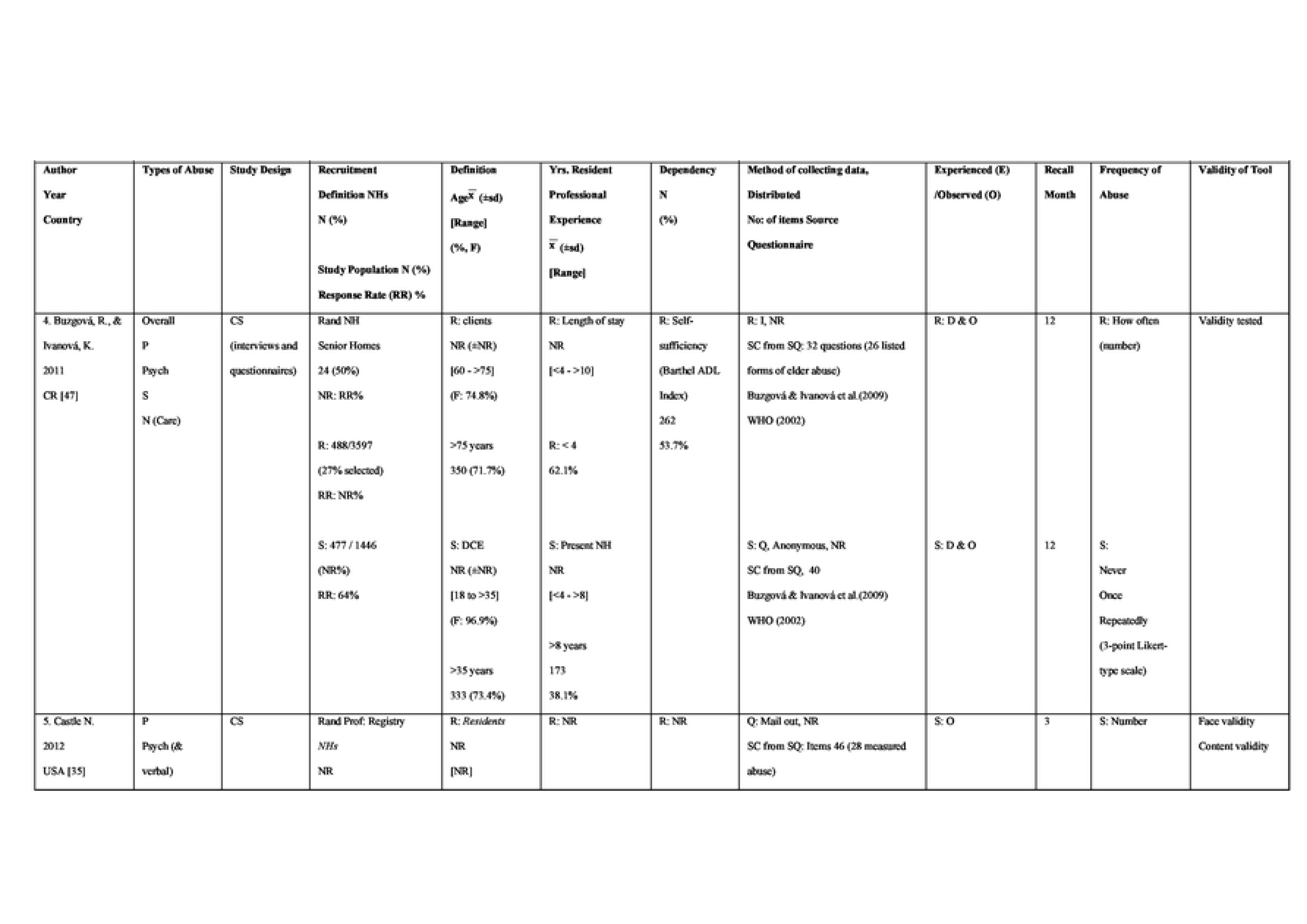
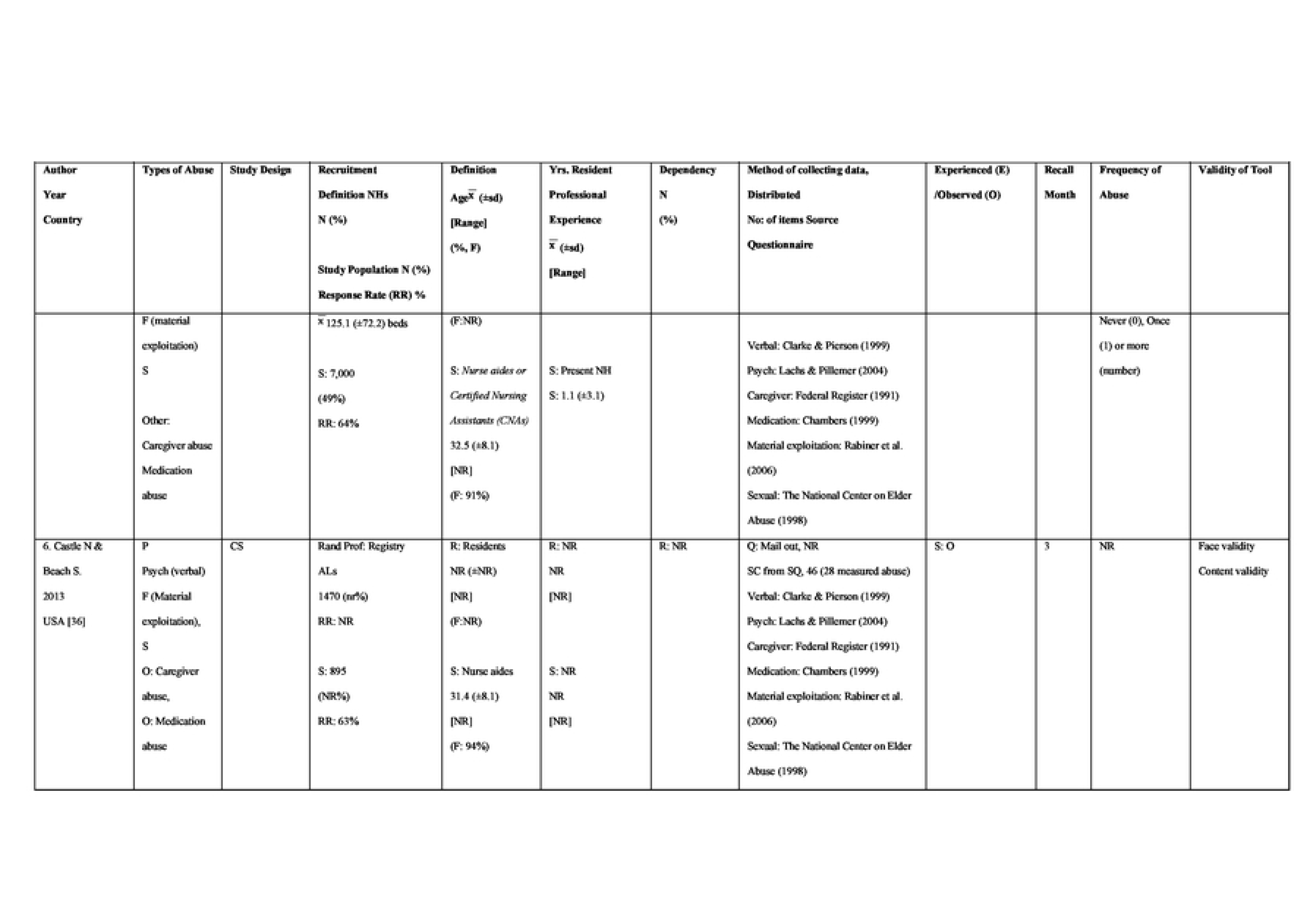
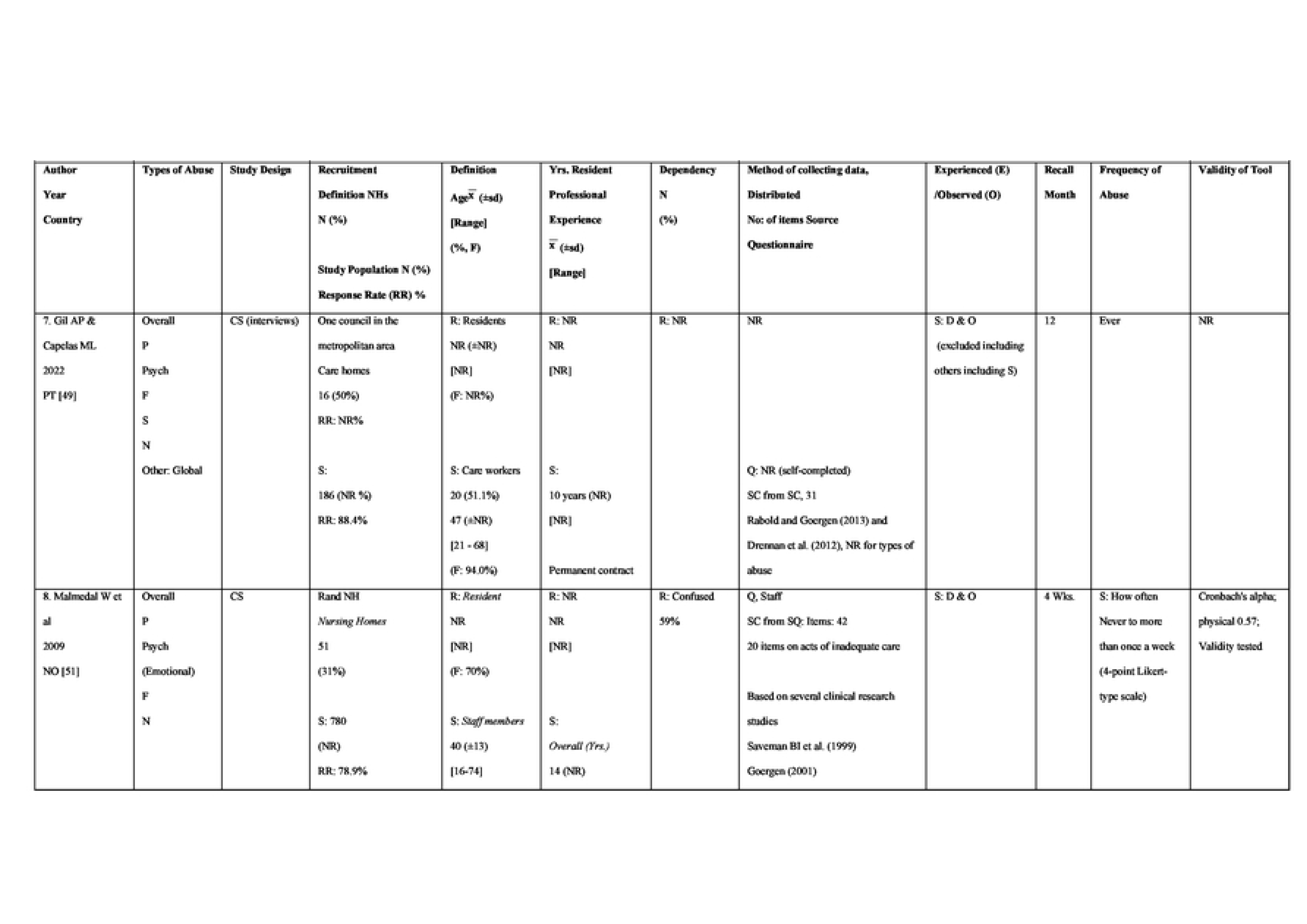
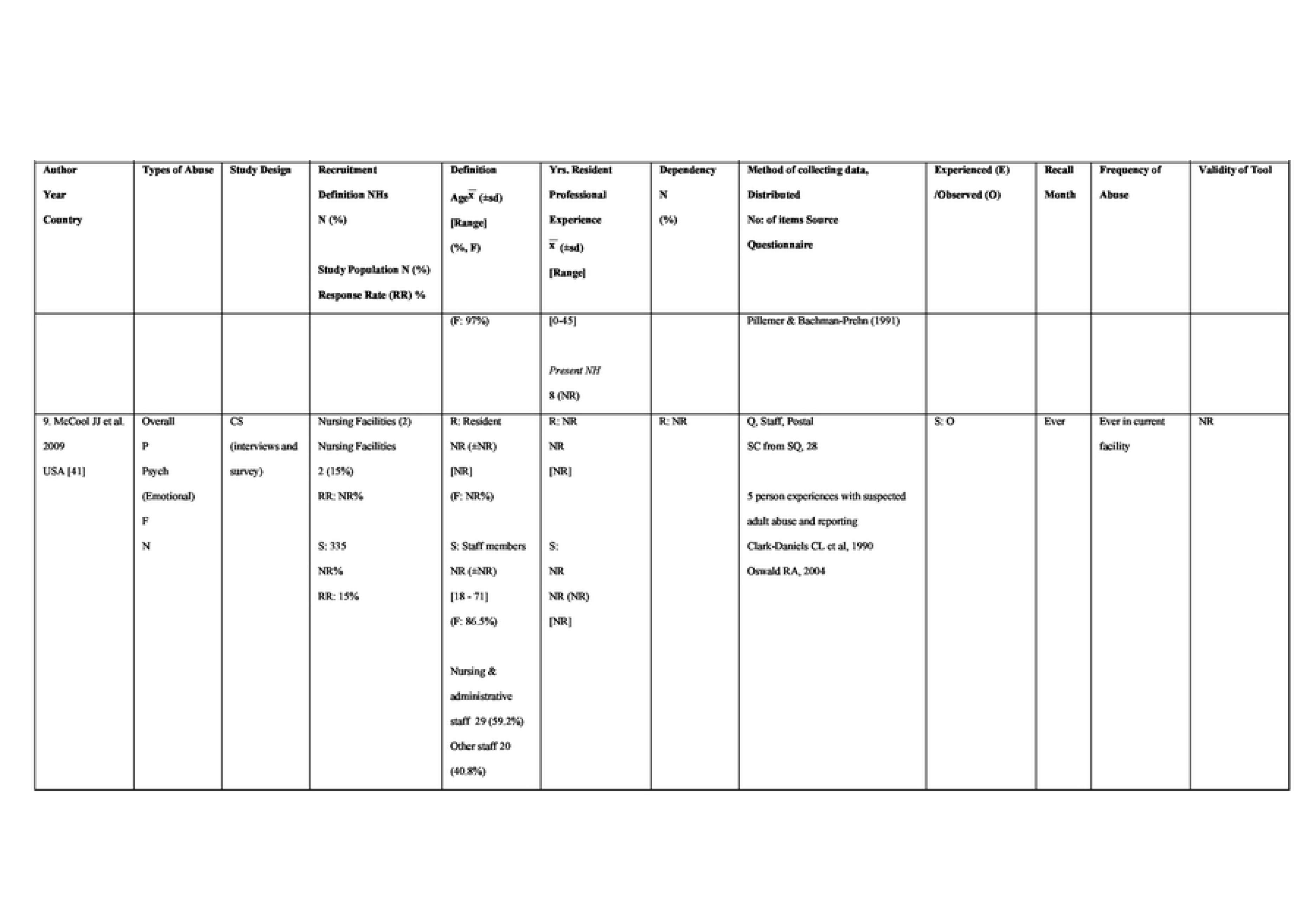
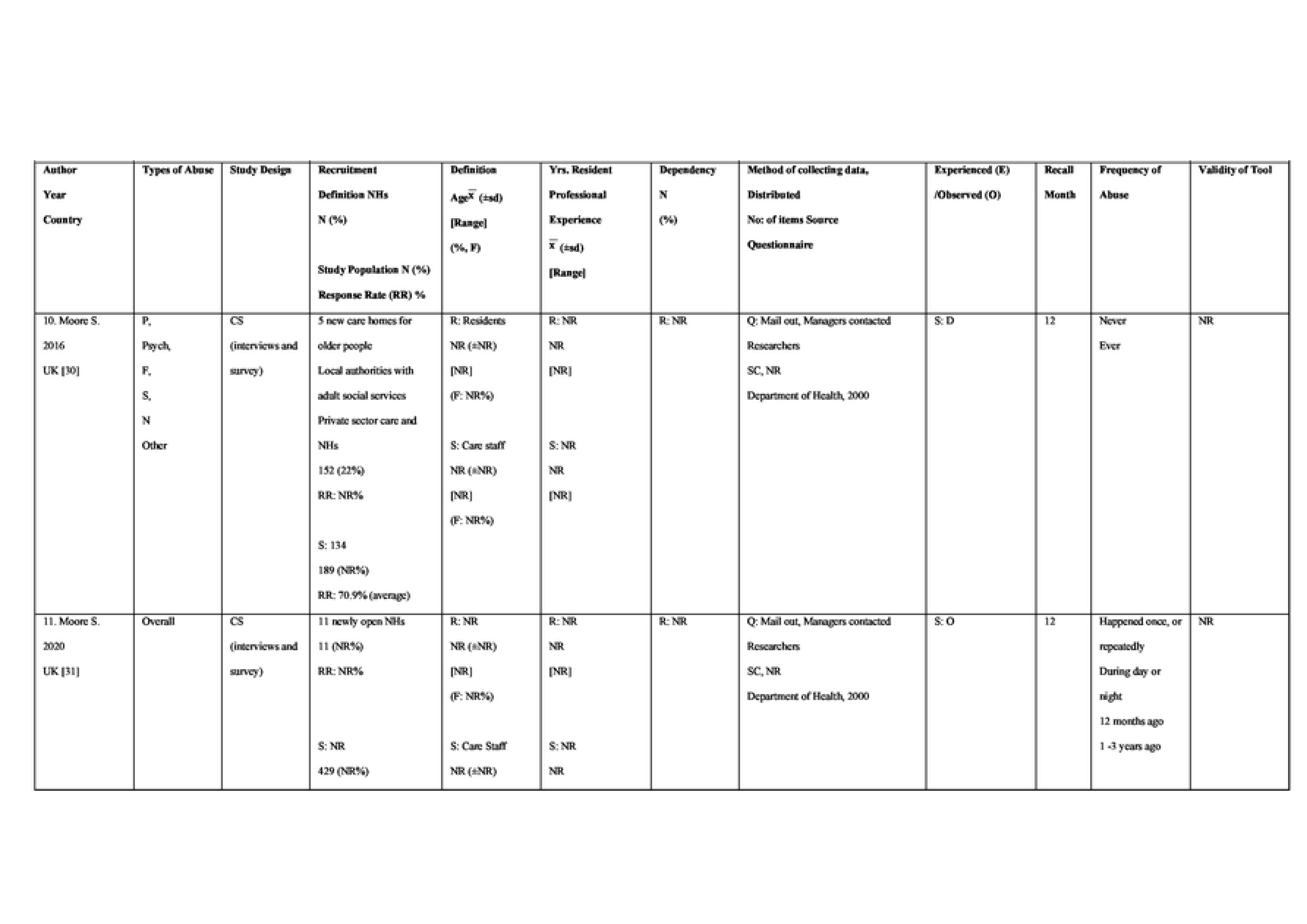
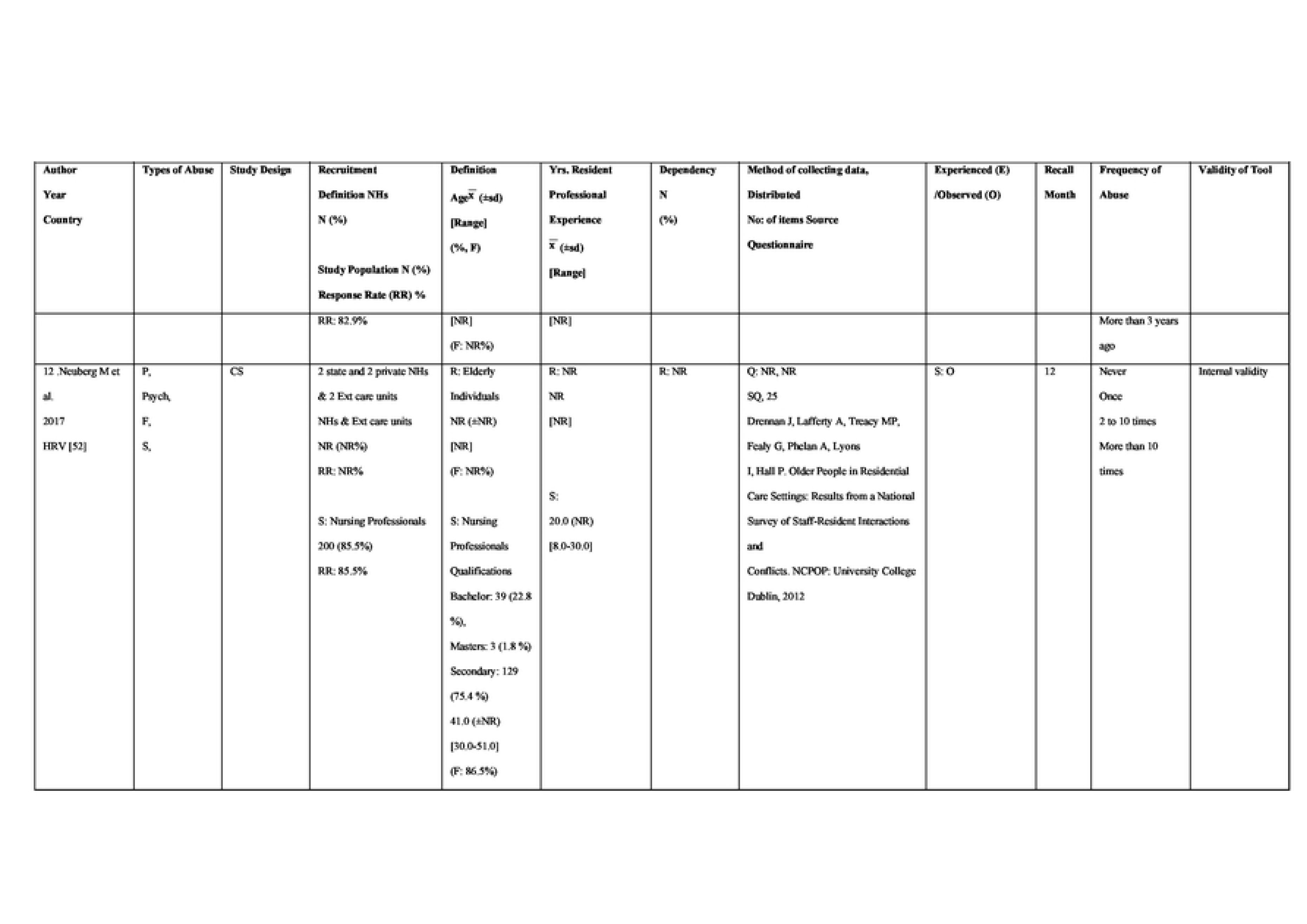
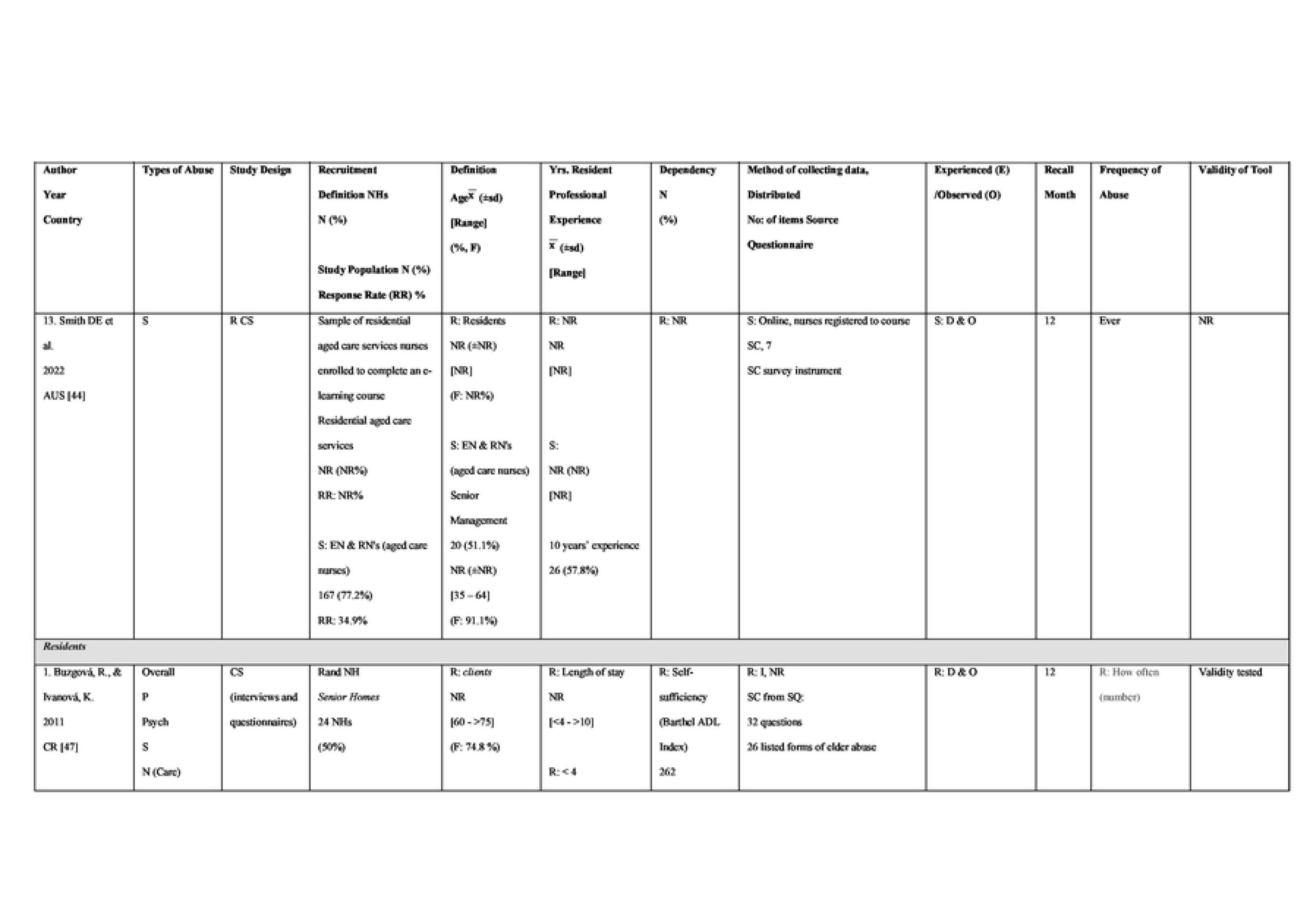
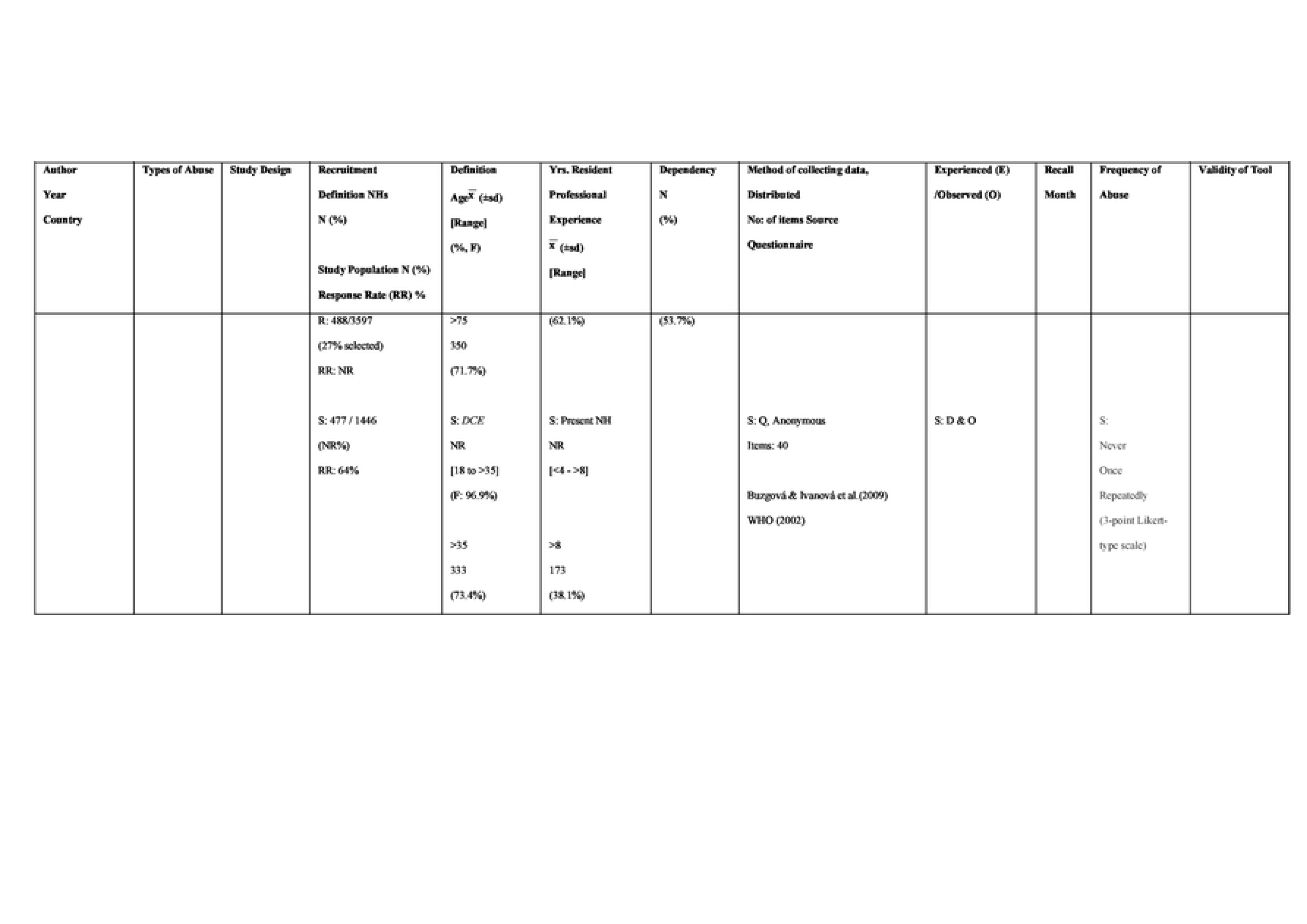
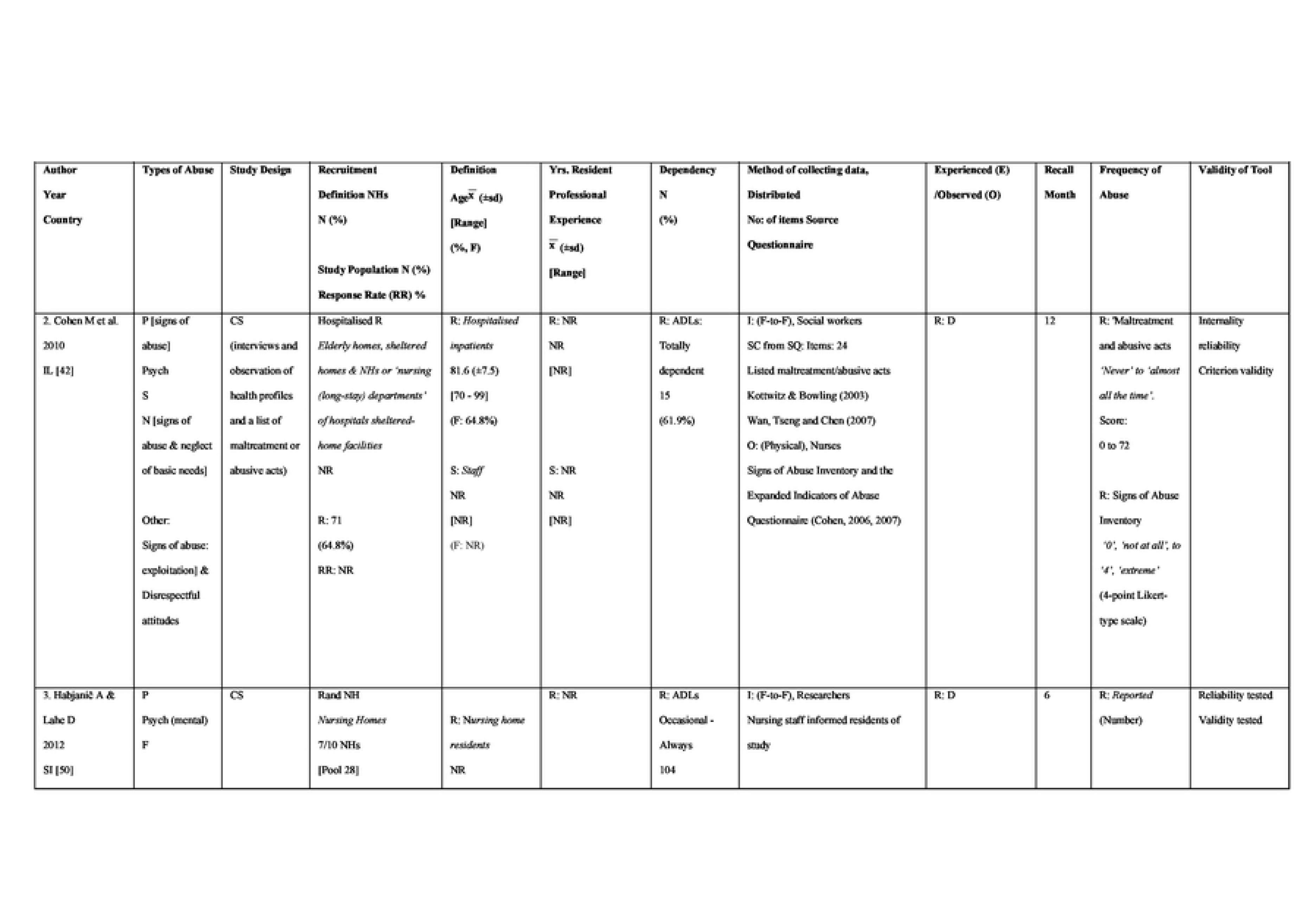
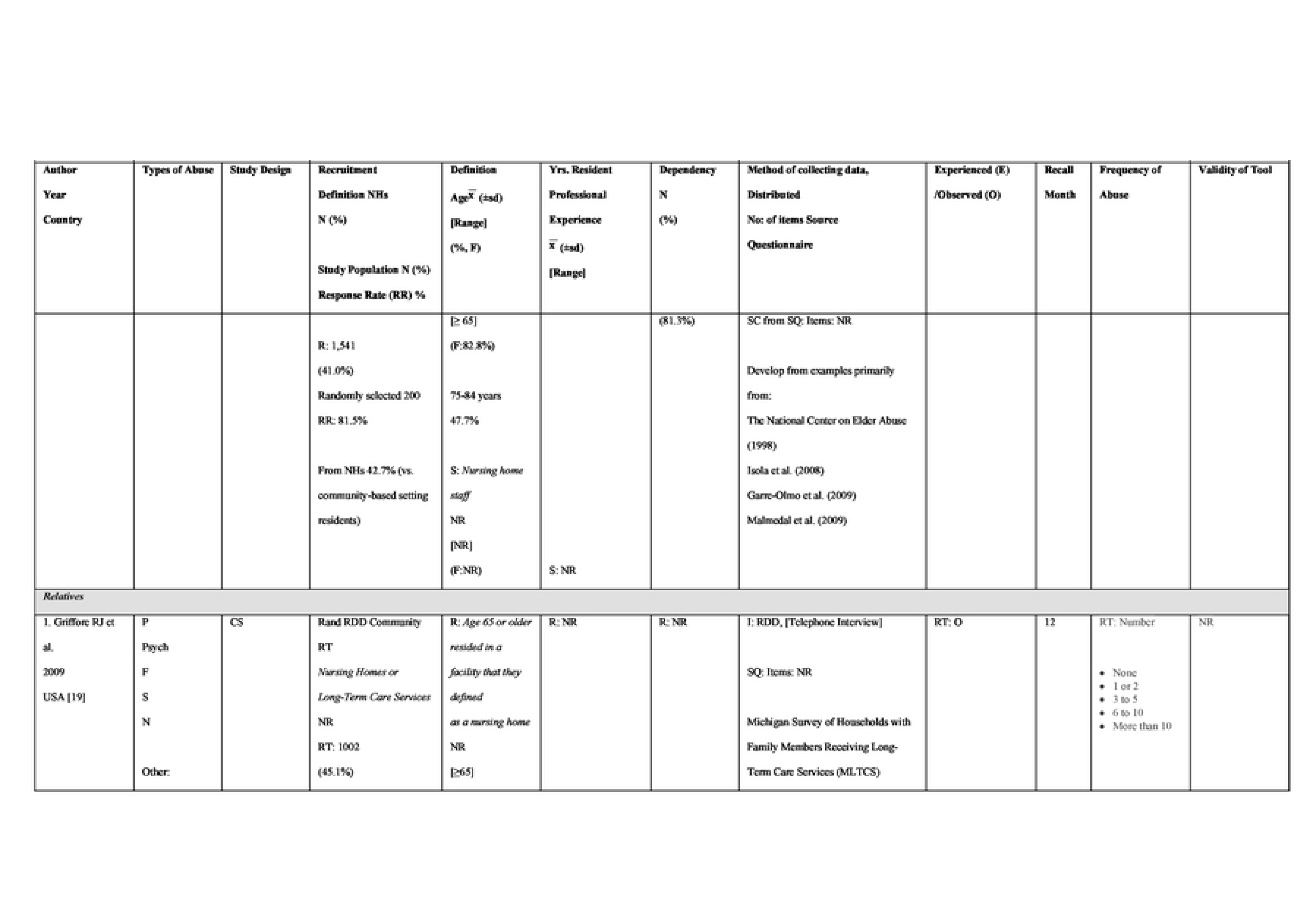
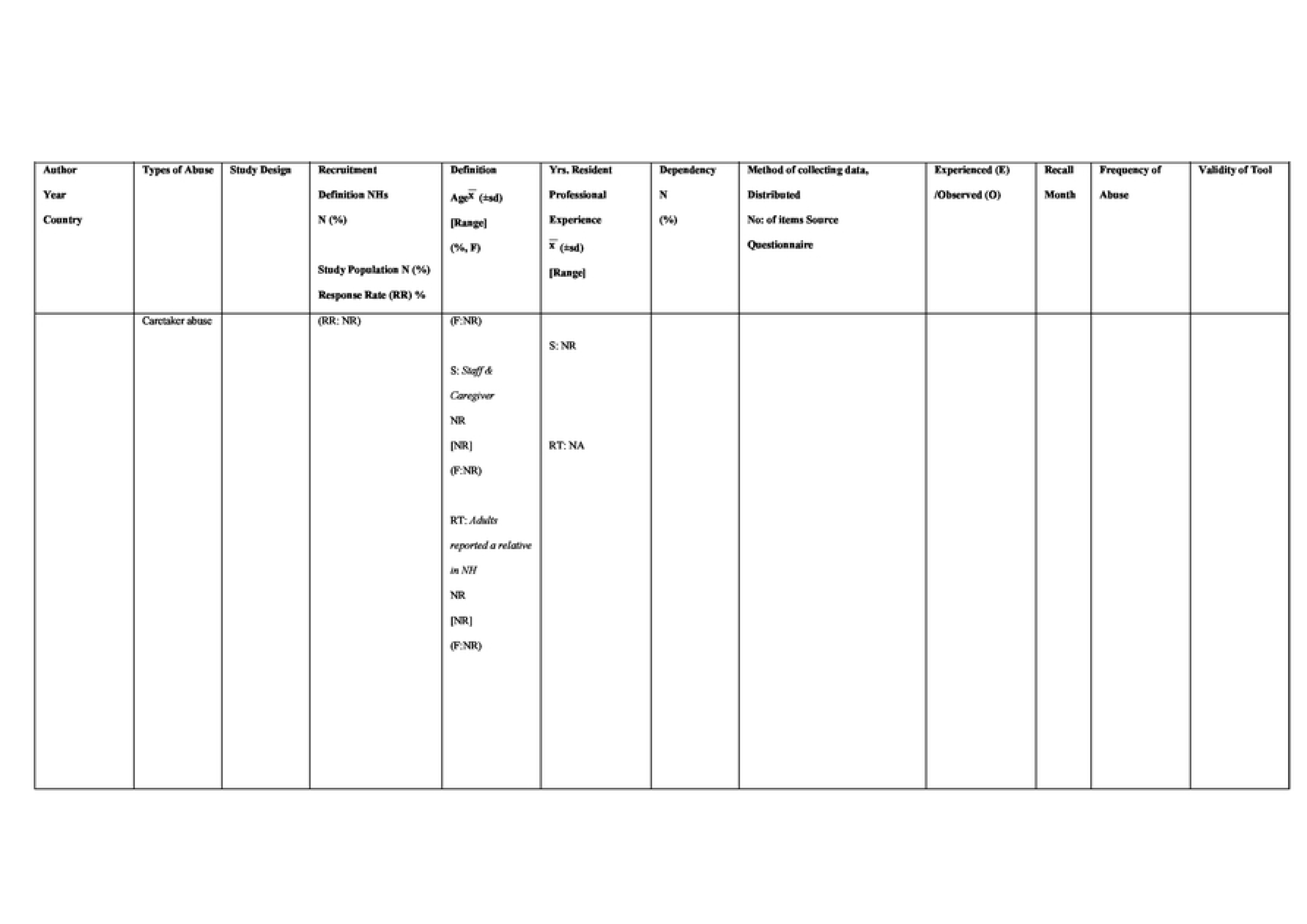
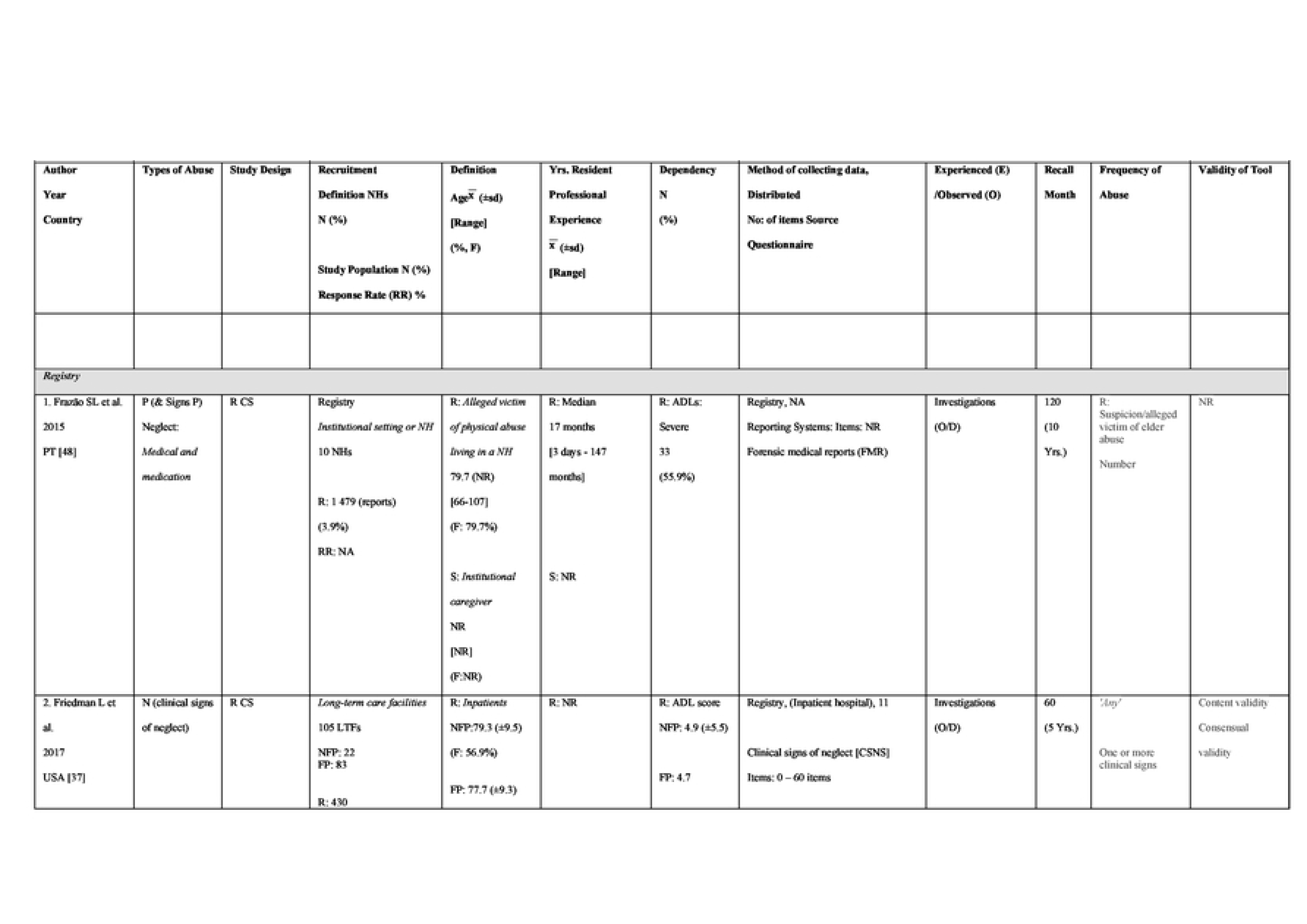
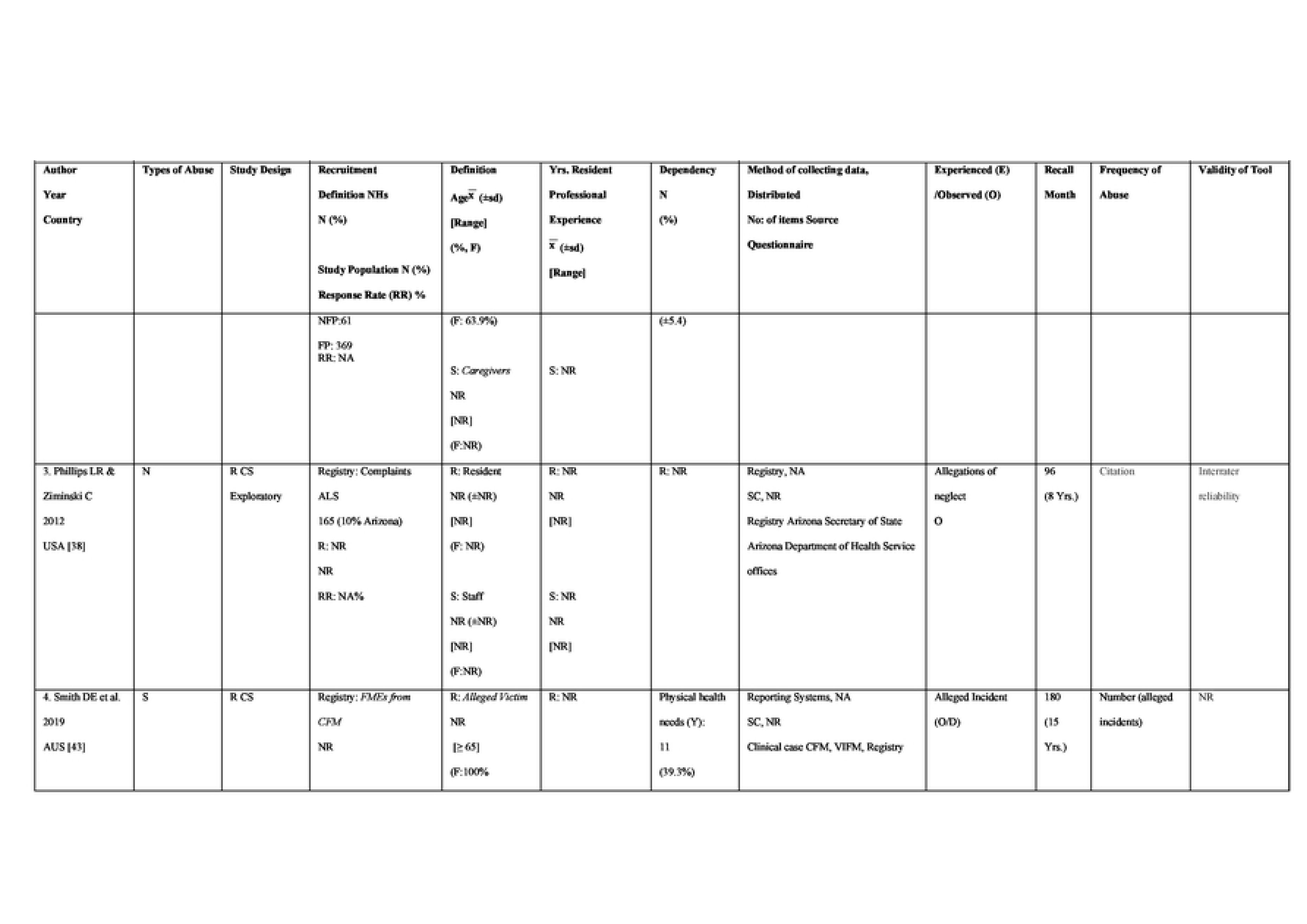
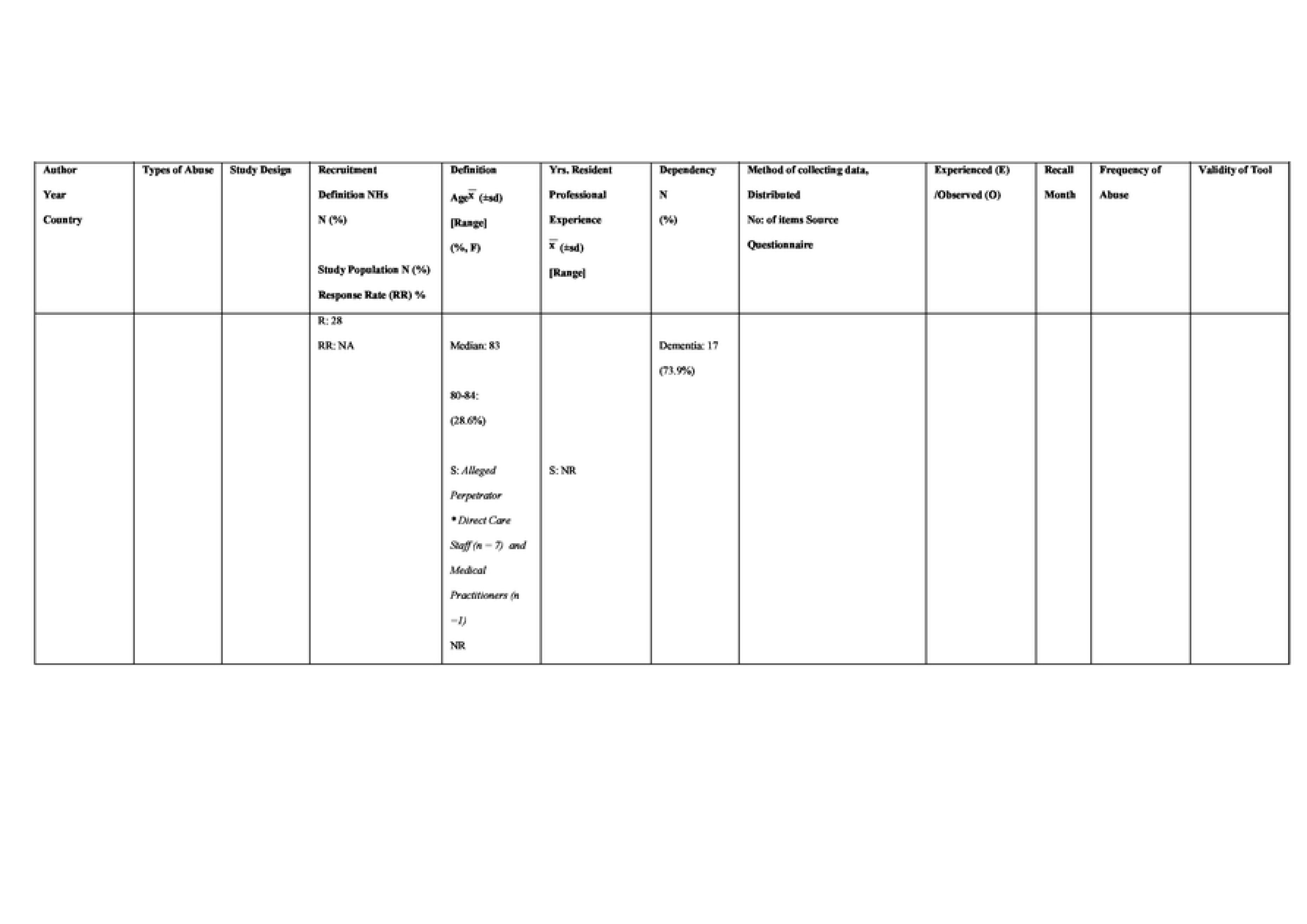
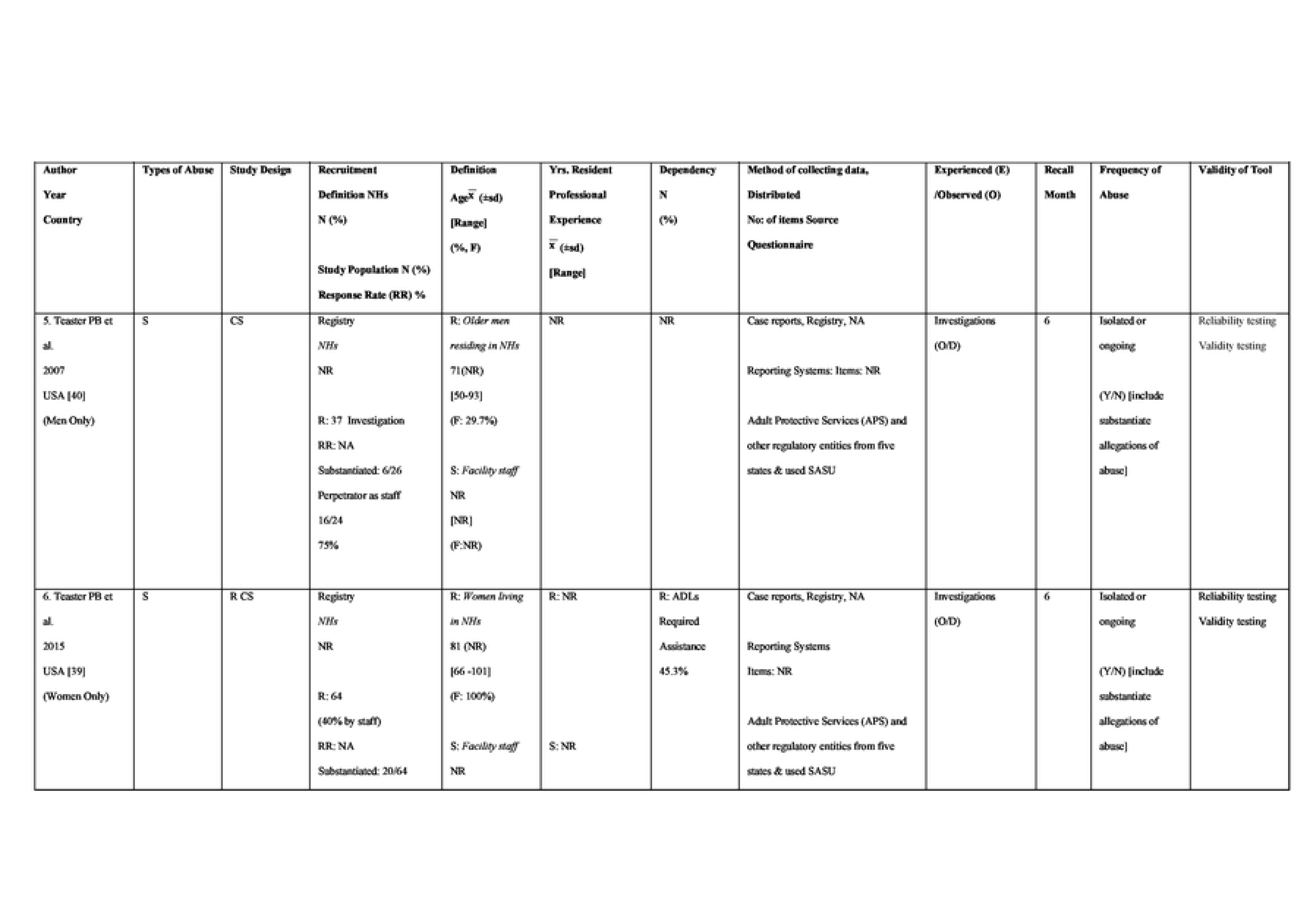
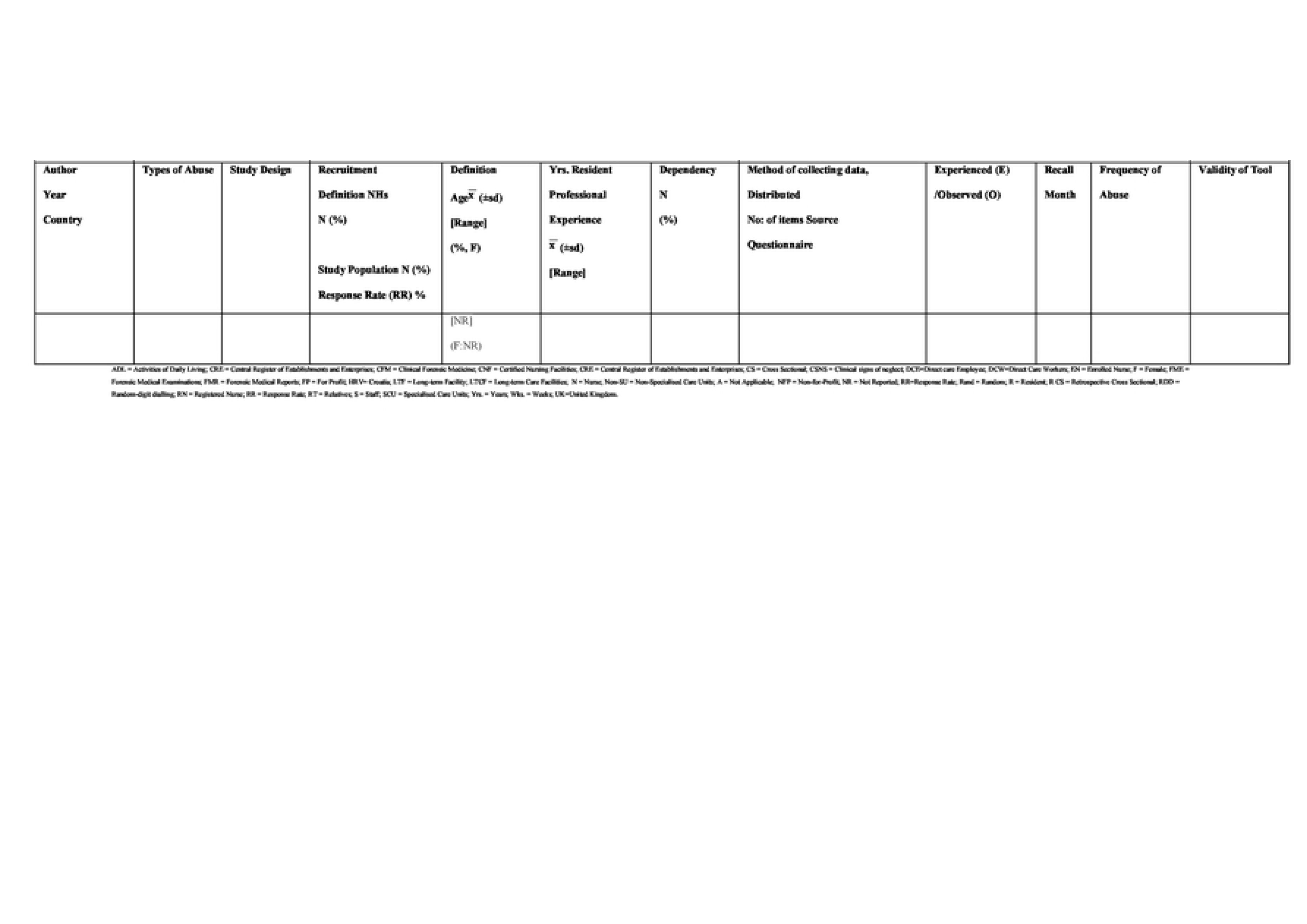
Summary of study characteristicsvia staff, residents, relative and community reporting abuse.

### Methodological Quality Assessment

A methodological quality assessment of included articles were independently assessed by three reviewers (TM, MA, and IK) using Boyle et al.(1988) [18] 8-item checklist, designed to evaluate the elements of prevalence studies (S4 File).

## Results

### Systematic review

A total of 1,298 citations were retrieved from the search. Four additional articles were located through hand searching. Duplicates and non-English language papers were then removed resulting in 826 records. Initial screening, against inclusion criteria, of title and abstract, reduced the records to 108. Detailed screening, through full-text review, reduced the records to 44 articles identified as meeting the study criteria. Four papers, by Griffore et al. (2009) [19], Page et al. (2009) [20], Post (2010) [21], Schiamberg et al. (2012) [22] and Zhang et al. (2011) [23], all reported data from the same study population. Griffore et al. (2009) [19] was subsequently retained over the other three, based on a stronger study design including a more defined recall period and a focus on multiple types of abuse. Papers published by Ben Natan et al.(2010) [24, 25] and Moore (4) [26-29] used the same population. Ben Natan et al.[25] study examining psycho-social factors affecting elders’ maltreatment in long-term care facilities and Moore’s paper examining observed abuse from two time periods, 2011 to 2013 and from 2015 to 2019 with prevalence data were chosen [30, 31]. While other studies did not provide prevalence data of abuse [32, 33] or examined perception of elder abuse and neglect among nursing staff working in a hospital [34]. The final study cohort comprised 22 studies (Fig. 1: Identification and Selection of Studies).

**Fig. 1.**
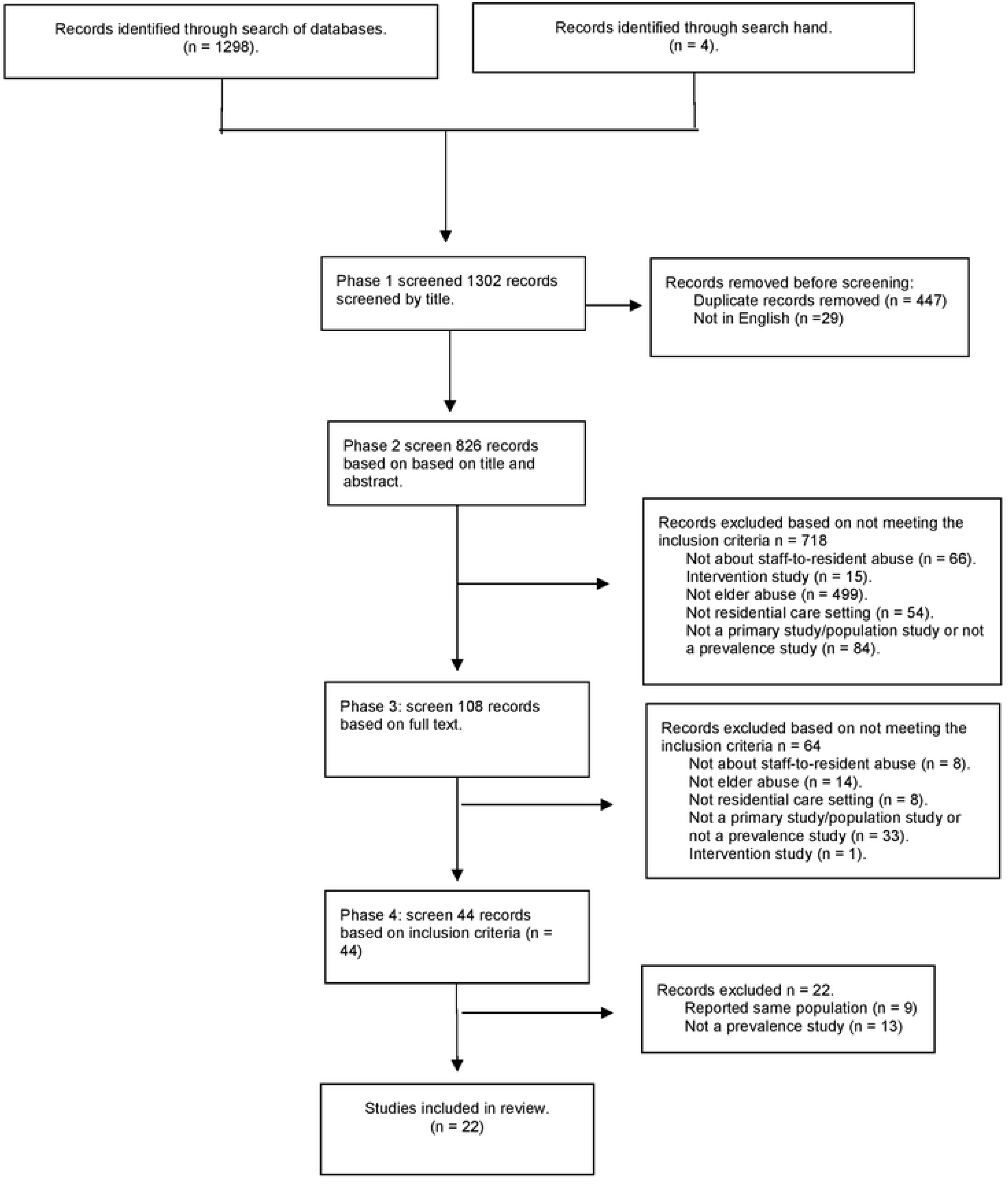
Identification and Selection of Studies - PRISMA flow diagram (14]

### Study Characteristics

Country settings varied, with eight from the United States of America [USA] [19, 35-41], two from Israel [24, 42], two from Australia [43, 44], nine from individual European countries [45-52] and two from the United Kingdom [30, 31].

Similarly, the studies were methodologically diverse, with 16 cross-sectional, 13 studies collected abuse data from staff, with the majority using surveys [35, 36, 44, 46, 47, 52] or questionnaires [24, 45, 49, 51]. Three studies utilised a mixed methods approach to distribute a staff questionnaire and interview [30, 31, 41], however for the purpose of this review we only included questionnaire data. A total of three studies were reviewed which collected data from residents. Residents were interviewed in two studies [47, 50], while one study by Cohen et al.(2010) [42], interviewed and collected data using participant’s health profile to determine signs of abuse from risk indicators of maltreatment or abusive acts.

The remaining cross-sectional study randomly selected ‘family members’ from the general community to participate in a telephone interview [19] and one from a community registry [40]. The five retrospective cross-sectional studies used one or more existing abuse reporting systems or registries [37-39, 48]. The following study characteristics are arranged based on persons reported abuse, from highest contact with resident (staff) to least contact with resident (registries) to examine recruitment methodology and study participant details (Table 1).

### Recruitment and Participant Characteristics – Staff

The 13 studies that recruited staff [24, 30, 31, 35, 36, 41, 44–47, 49, 51, 52] did so by selecting institutes or homes ranging from 2 [41] to 1,600 [45]. The majority of studies (7) utilised a randomised facilities sample of ‘nursing homes’ [45, 46, 51] including ‘extended care units’ [52] or ‘nursing facilities ranging from assisted living and independent living apartments’ [41], while two ‘long term institutionalized facilities’ [24], ‘senior homes’ [47]. Three studies utilised convenience sampling from ‘for profit’ [30] or/and ‘non-profit care homes’ [49] or based on the criteria a ‘newly open care facility’ [31]. All studies selected facilities within a defined geographical region. The remaining three studies recruited staff using a professional registry of Certified Nursing Assistants [CNAs], nurse aids within a state in the USA [35, 36], or from a subscribers list from a quarterly electronic resource addressing resident safety [44].

Staff were defined as ‘direct care workers’ or ‘employees’ [45, 47],’ care workers’ [49] or ‘care staff’ [30, 31]. Others defined staff as ‘facilities employees’ [41]. Some studies used a generalised staff in terms such as ‘staff workers’ [24], ‘staff members’ [51]. The six studies distinguished nursing professions as specific either ‘enrolled’, ‘registered nurses’ and/or ‘nurse aids’ or ‘certified nursing assistants’ [CNAS] [35, 36, 41, 43, 46, 52]. Of the 13 studies recruiting staff, 11 reported on one or more staff characteristics from gender to years of employment (see Table 1) [24, 35, 36, 41, 44–47, 49, 51, 52]. Of those who recruited staff cohorts ranged from 53 [44] to 7,000 [35, 36].

Of the 13 studies, 12 studies reported a response rate, ranging from 15% [41] to 92% [24] (Table 1). Staff participant characteristics were reported in eleven studies; ten reported the majority being female (>82%) [24, 35, 36, 41, 44–47, 49, 51, 52]; with an age range between 16 to 74 in seven studies, [51] and 13.7 mean years experienced as reported in five studies [24, 44, 49, 51, 52]. Current nursing home experience ranging from 1.1 [35] to 8 years [51], while one study reported the overall professional experience as a mode, with approximately 40% having more than 8 years’ experience [47]. Of the thirteen studies that utilized reported abuse by staff, four collect data on one or more resident characteristics [41, 45, 47, 51](Table 1).

### Recruitment and Participant Characteristics – Residents

Three studies examined older adult abuse as reported by residents [42, 47, 50]. One study selected 10 ‘nursing homes’ with 640 eligible ‘nursing home residents’ invited 200 to participate, with 82% doing so [50]. A second study selected 24 ‘senior homes’, screened 1,807 ‘clients’, with 27% meeting the study criteria [47]. No response rates were recorded. The third study collected data from 71 admitted ‘inpatients’ from ‘elderly homes’ or ‘nursing homes’ [42]. Studies examining residents were defined as a ‘client’ [47], ‘resident’ [19, 35, 45, 46, 50, 51] or a ‘patient’ [24, 37, 42](Table 1).

All three studies recruiting residents reported on residents’ characteristics. The majority of the participants were female (>65%) [42, 47, 50] with a mean age of 82 (±7.5) (range: 60 – 99) [42, 47, 50]. Activities of Daily Living (ADL) was used in several studies to report resident dependency levels. ADL were described as a frequency (from “from never to occasionally or always, needs help or assistance with all ADL’s), with the highest being ‘occasional to always’ (81%) [50], as a category, ‘required assistance with all ADLs’ (45%) [42]or as a status, ‘being self-sufficient’ (54%) [47]. Years of residency were only reported in one study as ‘length of stay’, with two-thirds reporting less than four years [47]. No staff characteristics were reported in the studies recruiting residents (Table 1).

### Recruitment and Participant Characteristics – Relatives

Relatives were recruited in only one study. There were 450 participants from the general community who had ‘a family member’ ‘receiving long-term care services’ to take part in a telephone survey [19]. No response rate was recorded. The only study characteristics collected were description of residents, as the ‘family member’ being female (73%). No staff characteristics were reported (Table 1).

### Recruitment and Participant Characteristics – Registry

Six studies utilized existing registries to report institutional abuse. Four studies from the USA, with two recorded close to 100 ‘sexual abuse’ reports reported in ‘certified nursing facilities’ over a short period of six-months [39, 40], while the third studies examined 616 complaints related to ‘neglect’ from ‘assisted living facilities’ over an eight-year period [38]. Lastly the final study examined ‘430 residents living in long-term care facilities’ admitted to one of the five metropolitan hospitals (as an inpatient) over a five-year period [37].

The two remaining studies from Australia [43] and Portugal [48], examined forensic medical reports for incidences of ‘abuse’ [48] or ‘female sexual abuse’ [43] from ‘nursing homes’ or ‘institutional settings’ over a 10 [48] to 15 [43] year period resulting in small prevalent cohorts, ranging from 28 [43] to 59 [48], respectively (Table 1). Residents were defined as ‘living or residing in nursing homes’ or as an ‘alleged victim’ [43, 48].

Of the six studies, five reported the percentage of female residents ranging from 58% [37] - 100% [39], with the exception of one study which focused on male residents only [40]. In these five studies, mean age was reported in three studies, with an average 79.7 years [37, 39, 48], while two studies reported on median age, ranging from 71 – 83 [39, 40, 43]. There was variation in defining resident’s dependency. Using the Activities of Daily Living (ADLs), dependency was reported as a status [‘being mildly to totally dependent’(62%)] [39], a category [‘mild/moderated’ or ‘severe’ (‘loss of autonomy’ [highest, severe 56%])] [48], a gradient [‘ability to function independently’ 0 – 10 scale (aggregate mean 4.8 [Barthel Index])] [37] or as a status [‘dependency or assistance with ADLs’ (18%)] [43]. Years of residency was only reported in one study, at the time of registered abuse, with a median of 17 months [48] (Table 1).

### Methodology Instruments Used to Measure Older Aged Abuse

Overall, we identified 16 instruments used to measure older aged abuse in long term institutes over the last 18 years. In Table 1, we constructed columns examining the methodological approaches for each of the studies included in the review, based on who reported the abuse (highest contact with resident, staff to lowest registries) with the following methodological features such as 1) method of administering data (such as mode, distribution and collection) 2) number of items and name of the identified instruments [including source], 3) observed and/or experienced abuse with type of abuse reported 4) recall period and 5) validity of tool. In S5 File, we have provided a more in-depth analysis of the commonly defined types of abuse (as defined by WHO), with additional information such as name of tool, source of tool, how type of abuse is defined and definition of type of abuse to examine homogeneity in each tool when measuring type of abuse.

### Instruments Used to Measure Older Age Abuse - As Reported by Staff, and /or Residents

Of the 16 instruments, 11 were used to measure abuse as observed and/or experienced by the staff member, resident or relative. The review found the three most commonly used tools measured staff abuse; Malmedal et al’s 42 acts of inadequate care instrument (2009) [45, 50, 51]; Castle’s (2012) 28 item questionnaire measuring how often staff observed and/or perpetrated abuse [35, 36, 46] and Drennan et al.(2012) [49, 52, 53] national survey on interactions and conflicts within nursing home settings. There were variations among these instruments ranging from modes of delivery, either presented as a questionnaire or survey, with differences in definition and types of abuse and discrepancies in recall periods.

Malmedal’s et al.(2009) [51] original 42-item questionnaire was used to measure staff ‘observed’ or ‘committed’ ‘physical, psychological (emotional), financial and neglect’ acts (unintentional and intentional acts) of inadequate care’ within a four-week recall period, using a four-point Likert-type frequency scale ranging from ‘never’ to ‘more than once a week’ was used in two other studies. This questionnaire has been tested for face validity only, indicating the tool was easy to follow and comprehensive, evidence pertaining to the other items of validity were not reported.

In 2012, Habjanič and Lahe [50] further modified Malmedal (2009) [51] question which asked ‘residents’ face-to-face if they had ‘ever experienced’ ‘mental’, ‘physical’ and ‘financial abuse’ using a ‘six months’ recall period to record the ‘number of incidences’, rather than using the Malmedal et al.(2009) [51] Likert scale (Table 1). In 2017, Blumenfeld Arens et al.[45] used Malmedal et al.(2009) [51] to ask staff if they only ‘observed’ ‘elder abuse’ (not as the original definition, ‘inadequate acts’ of ‘physical, psychological (emotional) and neglect, but not financial abuse, using with the same recall period and Likert-type frequency scale. None of the studies measured correlation ecoefficiency.

Castle and Beach’s (2013) [36] 46-item questionnaire measured the ‘number’ of times staff ‘observed’ ‘physical’, ‘psychological (verbal)’, ‘financial (material exploitation)’ or ‘sexual abuse’ in ‘last three months of their prior place of employment’ and was used again in a study by Castle the following year [35]. The questionnaire has been tested for face and content validity (using Fleisch–Kinkaid Scale), indicating this tool is measuring the degree which abuse is measuring abuse accurately Recently, Botngård et al.(2020) [46] changed Castle’s questionnaire to examine staff’s incidences of not only ‘observed’ but also ‘perpetrated’ abuse using the same recall, measuring additional types of abuse, ‘overall’ and ‘neglect’, within their ‘current place of employment’ in the ‘last 12 months’, not their prior place of employment, as the original instrument intended use. All studies utilized the original frequency using a three-point Likert-type scale as ‘never’, ’once’ or ‘repeatedly’ (Table 1). None of the studies reported on correlation ecoefficiency.

Gil & Capelas (2022) [49], and Neuberg (2017) [52] utilised the long-established questionnaire by Drennan et al.(2012) [53], a 25-item national survey of staff-resident interactions and conflicts within residential care settings. Between the two papers, there were variations with types of abuse and whether it measured as witnessed [52] and/or committed [49] abuse. In Neuberg et al. [52] study, the survey was pretested in a validation pilot study and achieved a reliability coefficient was > 0.7, deeming the instrument to be reliable.

One study utilised as part of their questionnaire, the long-established Iowa Dependent Adult Abuse Nursing Home Questionnaire [54] to measure ‘number of times’ ‘perpetrating’ or’ witnessing’ ‘acts of violence’ (including physical, psychological, financial [exploitation], sexual acts and neglect) ‘committed’ by staff ‘in the past 12 months’. This questionnaire was tested for reliability (0.89) [24]. The authors of this instrument have conducted readability and content validity analysis, although this was conducted in 2005 [55].

The remaining studies constructed questionnaires, surveys or telephone interviews using peer-reviewed studies and/or industry reports [19, 30, 31, 41, 42, 44, 47, 49] to develop their own surveys with 7 – 40 items [44, 47], to report abuse by staff, residents or relatives as experienced, suspected or observed, with no reporting on any validation studies being carried out. Overall, there is still some heterogeneity among these instruments, they are still in their early constructs, more studies and methodology testing are required conducted to validate these instruments. See Table 1 for further details.

### Instruments using Data Registries to measure Older Age Abuse

The remaining six studies utilized government registries or databases. Four studies utilized existing government registries such as the Registry Arizona Secretary of State & Arizona Department of Health Service offices [38] or the Adult Protective Services (APS) (National Adult Protective Services Association [NAPSA], 2021) (3) in conjunction with a survey (Sex Abuse Survey [SASU]) [39] and/or with hospital records with the use of Clinical Signs of Neglect Scale (CSNS) [37, 39, 40]to report ‘isolated or ongoing’ investigation of ‘citations and allegations’ [38] or a ‘suspected, reported, unsatisfactory, partial or substantiated resolution case of abuse’ [39, 43, 48] or used the to identify ‘clinical signs of elder mistreatment or elder neglect’ [37]. Two studies utilized clinical forensic medicine reports [43, 48] of ‘current or past medical observations and/or victim complaints of suspicion of physical or psychological abuse’ [48] or ‘alleged incidence of sexual assault among women only’ to report incidences of abuse [43]. These studies varied with recall periods ranging from six months [39, 40]to 15 years [43]. Some of these studies due to governance process of reporting incidence cases into a registry required validated professional staff to perform examinations [48], a consortium of experts to develop clinical validated scales [37, 39, 40] and independent research reviewers [43] ensuring embedded testing for interrater reliability, validity and reliability of findings, however registers are commonly known for their practical limitations such as variability in data collected impacting quality (completeness or accuracy), difficulties with follow-up and data dredging (Table 1).

### Impact of Methodology on the Results

#### All abuse

Out of the 22 studies, ten studies measured the overall incidence of abuse (measuring one or more types of abuse as defined by WHO) [24, 30, 31, 41, 42, 46–49, 51](Table 1), with the highest overall prevalence reported over a four week period reported in one study, 91% ‘observed’ abuse by *staff*, while ‘committed’ abuse by staff was at 87% [51]. Two studies reported abuse by staff over a 12-month period resulted in lower rates of ‘observed’ abuse ranging from 55% [49] to 76% [46] and for ‘perpetrating’, from 54% [24]to 60% [46].

Two studies reported by ‘*residents’* overall abuse over a 12-month period, retained lower rates, than above. ‘Experienced’ abuse ranged from 11% [47]to 31% [42], while ‘observed’ was at a lower rate of 5% in one study [47]. No studies examined overall abuse reported by relatives or community via a registry. Five studies (23%) reported all five types of abuse as defined by WHO [19, 24, 42, 46, 49] (See Table 1). The following sections will examine prevalence based on types of abuse as defined by WHO, physical, psychological, financial, sexual and neglect.

#### Physical abuse

The most commonly measured form of abuse was physical abuse, also defined as ‘physical violence’ [24], ‘mistreatment [19], ‘maltreatment’ [42] or ‘acts of physical character’ [51], measured in 15 studies [19, 24, 30, 35, 36, 41, 42, 45-52]. An accumulation of 81 items were identified to describe acts of physical abuse, with each study using three [45] to 11 [42] items to describe physical abuse. The most commonly used verbs to describe physical abuse was ‘hitting’ (8) [19, 35, 42, 46, 47, 50] or ‘kicked’ (7) [19, 35, 46, 47, 50], with variations in definition, recall periods and persons reported. One study relied on staff to define physical abuse [30] or did not disclose items measured [31] (S5 File).

The highest rate of physical abuse reported was ‘witnessed’ by *staff* (44%), in the act of ‘restraining/hold back a resident’ ‘over a recall period of four weeks’ [51], and the highest ‘committed’ abuse was 33% from the same act of ‘restraining/hold back a resident’ as reported in the same study. When the same questionnaire was used in Blumenfeld Arens et al. [45] inn 2017, the study questionnaire, measured witness physical abuse over a 4-week period, resulting in a lower rate of 1.4%.

Studies examining physical abuse over a 12-month period, Gil and Capelas (2022) [49] and Neuberg et al. (2017) [52] using the same questionnaire [53], resulted in different levels of physical abuse by *staff.* Neuberg et al. (2017) [52] reported over 12 months, 42% of staff observed ‘force feeding the resident’ in the last 12 months, whereas Gils and Capelas (2022) [49] recorded 14% observed staff committing ‘at least 1 of the 6 behaviours of physical abuse’. The remaining studies utilised various measurement tools, with the same recall period of 12-months resulting in observed rates ranging from 6% [30] to 30% [47], while committed abuse were even lower ranging from 1.7% [30] to 12.3% [24].

Three studies reported physical abuse reported by *residents* either in the last six [50] to 12 months [42] resulted in lower rates of ‘observed’ abuse, from 1% [47] to 2% [47] and 8% for ‘experienced’ abuse [42]. Cohen’s study [42] found only three residents attained a score of three or more on the signs of physical scale. Compared to this, *relatives* reporting abuse in telephone interviews in the last 12 months had higher rates of abuse at 74% [19], while physical signs or evidence of physical abuse from forensic medical reports (FMR) from registries, were lower at 55 cases over a 10-year period [48], however these tend to be extreme cases of abuse (S5 File).

#### Psychological abuse

The second most common measured form of older age abuse in long term institutes was psychological abuse. Fourteen studies [24, 30, 35, 36, 41, 42, 45–47, 49-52] addressed psychological abuse. Three studies defined this type of abuse as ‘psychological abuse’ [30, 42, 46, 47, 49] while the remaining six defined as ‘emotional’ [19, 41, 45, 50], ‘mental abuse’ [24, 51] or as a combination of ‘psychological and verbal abuse’ [35] or ‘emotional or psychological and verbal mistreatment’ [19].

There were in total 47 items, with each study using three [45]to 14 [47] items to classify psychological abuse. The most common terms used to describe psychological abuse were of ‘intent’(5) [47] or ‘threat’(4) [35, 47, 50, 51]. A total of four studies did not disclose items or descriptions of types or examples of abuse asked [24, 41, 52] (S5 File).

Psychological abuse was reported by *staff* (11) [24, 30, 35, 36, 41, 45–47, 49, 51, 52], *residents* (2) [42, 50] and *relatives* (1) [19]. The highest rates of ‘observed’ and ‘committed’ act of psychological abuse was ‘entering a room without knocking’ of abuse by *staff,* 64% of staff committed the act, while 84% observed other staff in the last four weeks [51]. Three studies examine psychological abuse ‘committed’ by staff over the last 12 months found higher incidents ranging from 23% [24] to 46% [47], with variation in instruments utilized to measure this form of abuse. ‘Observed’ abuse by *staff* was reported in five studies, with incidents ranged from 30% [47] to 62% [19], with variation in instruments used making it difficult to provide an average rate. Two studies utilised Dennan et al. [53] instrument however there was a 20% difference between the act of shouting at resident in anger [49] [33% [49] from 16 care home settings versus 55% from nursing home and extend care units [52]].

*Residents* reported ‘experienced’ psychological abuse ranged from 10% over a 12-month period [47] to 56% [50] over a six-month period, however reported ‘observed’ abuse was lower at 4% [47]. Uniquely, Cohen et al. (2010) [42] reported distribution of disclosed abuse and found “very low complaints for psychological abuse” (13%). Telephone interviews among family members *(relatives)* reported 84% ‘observed’ ‘verbal mistreatment’ by nursing staff ‘in the previous year’ [19]. No studies measuring psychological abuse used registries. All studies examined specific psychological acts, making it difficult to aggregate the incident rate due to variations as shown above (S5 File).

#### Financial abuse

Eleven studies defined financial abuse either as ‘material exploitation’ [19, 35, 36, 46] and/or ‘financial exploitation’ [24, 42], ‘financial abuse’ [30, 49, 50], ‘acts of financial character’ [19, 46, 51]. An accumulation of 24 items was identified to described acts of financial abuse, with each study using one [51] to seven [42] items to describe financial abuse. Most common term used to describe financial abuse were ‘signing documents’ (6) [35, 42, 46, 50] (S5 File).

Most of the studies examining the rates of financial abuse were reported by *staff* (8) [24, 30, 35, 36, 41, 46, 49, 51], followed by *residents* (2) [42, 50]or *relatives* (1) [19]. The highest level of financial abuse reported in this review were observations of staff from *relatives* of older adults residing in nursing homes, 71.9% [19]. This was followed by reported ‘experienced’ financial abuse by *residents* in one study, at 32.8% [50] over the last 6 months. Lower rates of financial abuse were reported by staff, for ‘observed’ incident ranged from 2.1% [46] to 3.3% [49] in care and nursing homes.

S*taff* reporting ‘committing’ financial abuse were at a lower rate 0% [51] to <1% [24, 46] over the last four weeks to 12 months, while two study examined staff ‘observed’ financial abuse found 10%, of staff took ‘assets’ from nursing home residents or ‘destroying belongings’ of resident residing in assisted living institutes, 26% [36]. Interestingly Castle’s questionnaire used in two studies, in two similar setting and recall periods, found the incidents from ‘taking residents assets’ were similar, 10% [35] versus 11% [36]. No studies measuring financial abuse used registries (S5 File).

#### Sexual abuse

Eleven studies reported the prevalence of sexual abuse, described either as abuse [19, 24, 30, 36, 39–44, 46, 47, 49] with variation in definition of this form of abuse ranging from as an act of ‘assault’ [43], ‘misconduct’ [19], ‘violence’ [24], ‘unlawful or unwelcome sexual behaviour’[44] or ‘sexual nature without consent’ [49], to an outcome of signs of ‘forensic evidence’ [40] or ‘victimization (women)’ [39].

Number of items describing sexual abuse ranged from one [49] to 11 items [39]. Among the eleven studies, in total were 34 items that identified abuse a including, as an act of exposure to (4) [35, 39, 40] (hands off) to oral-genital contact (3) [35, 39, 40] (hands on). Evidence of signs of sexual abuse included a torn underwear to infected [42].

Most of the studies relied on reports by *staff* (4) [24, 35, 46, 47], or *registries* (3) [39, 40, 44], followed by direct reporting from residents (2) [42, 47]and one by *relatives* [19] (S5 File). Reports from *relatives* had the highest reported level of sexual abuse at 40% [19]. *Registries* reporting an incidence of sexual abuse performed by staff ranged from 15.6% to 25% [39, 40, 43] however these cases were over a ten-to-15-year period. The lowest report incidences of this type of abuse were reported by *staff* as ‘observed’ resulted in ≤ 7 % or ‘committed’ <1% [24, 30, 35, 36, 41, 46, 47, 49], while two studies reporting no sexual abuse reported by *residents* [42, 47]. There were two studies showing some consistency with findings, utilising the same questionnaire in different institutionalised settings, found staff observed 69 nursing home staff and 61 assisted living staff ‘exposed private body parts to embarrass resident’ in the last three months [35, 36] (S5 File).

#### Neglect

Similar to psychological abuse studies, neglect is the equally the second highest form of abuse investigated in this review among older adults residing in long term institutions [19, 24, 30, 37, 38, 41, 42, 45–49, 51, 52].

The definition of neglect varied with 3 [38] to 11 items [37] describing these acts from ‘physical and mental neglect’ [24], to ‘clinical sign of neglect’ [37, 42, 48], or collective categorised as ‘personal, environmental, medical’ [38] to specific items described care neglect such as ‘not changing the position of bedridden person’ or ‘ignoring resident when they called’ [47, 49, 52]. Only two studies utilised the same instrument to measure neglect [35, 36]. Four studies did not provide or specify items that were measured for this type of abuse [24, 41, 52].

Neglect ‘observed’ by *relatives* retain the highest rate in failure to provide basic needs to residents (86.9%) [19]. Four studies reported neglect ‘committed’ by *staff* over the last 12 months, with results varied from 1% [47] to 46.9% [46], compare to nine studies reporting ‘observed’ acts of neglect ranging from 9% [47] to 57.8% [46]. These variations are due to different instruments and definitions used to measure neglect. Surprisingly, four studies used the same instruments, however disseminated findings differently, with one study reporting if ‘observed’ or ‘committed’ one of the ten items list for neglect, while the other reported 10 items distinctly with respected incident rates [45, 49, 51, 52].

The highest prevalence of neglect was 24%, attained from the face-to-face interviews conducted by hospital staff [42] among inpatient *residents*, while the another study when interviewing residents on ‘observed’ or ‘experienced’ neglect conducted in facilities were ‘unmentioned’ [47]. *Registries* reported 20% of severe cases of neglect, however again, this was over a 10 or more-year period [37, 48], while other study reported a total of 1,196 total neglect allegations, with 535 substantiated, over an eight year period, making it difficult synthesis findings [38]. Other abuse items not classified by WHO are also included in S5 File table.

### Methodological Quality Assessments

Studies were assessed and ranked by methodological score and categorized according to their study design and sampling (Table 2); using an eight-item methodological scoring standardized checklist [18]. Two independent reviewers scored a total of 88 items and agreed on 82 (93%) (κ 0.90 (95% CI 0.88 to 0.99), p<0.001) meaning there was a high agreement. The only minor discrepancy was from the interpretation of validated measurement tools. The representation of samples was at times not reported (85%), with no studies examining non-respondents and five studies (36%) reporting response rates [24, 35, 46, 51]. Only one study accounted for sampling design in their analysis [45], while all studies did not report confident intervals for prevalence rates (item 8). A total of four studies (29%) achieved a total score above 5 [45, 46, 50, 51], with scores ranging from 1/8 to 6/8.

**Table 2:** Methodological Qualuy of Studies.

|  | Author, Year, Country | Mode of Recruitment | 1 | 2 | 3 | 4 | 5 | 6 | 7 | 8 | Total |
| --- | --- | --- | --- | --- | --- | --- | --- | --- | --- | --- | --- |
| **Number** | **NURSING HOMES** |  |  |  |  |  |  |  |  |  |  |
| 1. | Botngård A et al, 2020, NO [46] | NHs | 1 | 1 | 0 | 1 | 1 | 1 | 1 | 0 | 6 |
| 2. | Habjanič A & Lahe D, 2012, SI [50] | NHs | 1 | 1 | 1 | 1 | 1 | 1 | 0 | 0 | 6 |
| 3. | Blumenfeld Arens O, 2017, SW [45] | NHs | 1 | 1 | 0 | 1 | 0 | 1 | 1 | 0 | 5 |
| 4. | Malmedal W et al, 2009, NO [51] | NHs | 1 | 1 | 0 | 1 | 1 | 1 | 0 | 0 | 5 |
| 5. | Buzgová, R & Ivanová, K, 2011, CR [47] | NHs | 1 | 1 | 0 | 1 | 1 | 0 | 0 | 0 | 4 |
| 6. | Castle N, 2012, USA [35] | NHs | 1 | 1 | 0 | 1 | 1 | 0 | 0 | 0 | 4 |
| 7. | Ben Natan M et al, 2010, IL [24] | NHs | 0 | 1 | 1 | 1 | 0 | 0 | 0 | 0 | 3 |
| 8. | Griffore RJ et al, 2009, USA [19] | CATI | 1 | 1 | 0 | 1 | 0 | 0 | 0 | 0 | 3 |
| 9. | Neuberg M et al, 2017, HRV [52] | NHs & ECUs | 0 | 0 | 0 | 1 | 1 | 1 | 0 | 0 | 3 |
| 10. | Smith DE et al, 2022, AUS [44] | Registered subscribers to resource on resident safety | 1 | 0 | 0 | 1 | 0 | 0 | 0 | 0 | 2 |
| 11. | Gil AP & Capelas ML, 2022, PT [49] | NHs | 0 | 0 | 0 | 1 | 0 | 0 | 0 | 0 | 1 |
|  | **ASSISTANT LIVING** |  |  |  |  |  |  |  |  |  |  |
| 1. | Castle N & Beach S, 2013, USA [36] | Professional Registration Nurse Aides | 1 | 1 | 0 | 1 | 1 | 0 | 0 | 0 | 4 |
| 2. | McCool JJ et al, 2009, USA [41] | ALF & ECUs | 0 | 0 | 0 | 1 | 0 | 0 | 0 | 0 | 1 |

| **CARE HOMES** |  |  |  |  |  |  |  |  |  |  |  |
| --- | --- | --- | --- | --- | --- | --- | --- | --- | --- | --- | --- |
| 1. | Moore S, 2016, UK [30] | CHs | 0 | 0 | 0 | 1 | 0 | 0 | 0 | 0 | 1 |
| 2. | Moore S, 2020, UK [31] | CHs | 0 | 0 | 0 | 1 | 0 | 0 | 0 | 0 | 1 |
| **REGISTRIES** |  |  |  |  |  |  |  |  |  |  |  |
| 1. | Teaster PB et al, 2007, USA [40] | RG | 1 | 0 | 0 | 1 | 1 | 1 | 0 | 0 | 4 |
| 2. | Teaster PB et al, 2015, USA [39] | RG | 1 | 0 | 0 | 1 | 1 | 1 | 0 | 0 | 4 |
| 3. | Phillips LR & Ziminski C, 2012, USA [38] | RG from ALFs | 1 | 0 | 0 | 1 | 0 | 1 | 0 | 0 | 3 |
| 4. | Smith DE et al, 2019, AUS [43] | RG(s) | 1 | 0 | 0 | 1 | 0 | 0 | 0 | 0 | 2 |
| 5. | Frazão SL et al, 2015, PT [48] | RG | 1 | 0 | 0 | 0 | 0 | 0 | 0 | 0 | 1 |
| **HOSPITALS** |  |  |  |  |  |  |  |  |  |  |  |
| 1. | Cohen M et al, 2010, IL [42] | Hosp | 1 | 0 | 0 | 1 | 1 | 1 | 0 | 0 | 4 |
| 2 | Friedman L et al, 2017, USA [37] | Hosp | 1 | 1 | 0 | 1 | 1 | 0 | 0 | 0 | 4 |

ALFs=Assistant Living Facilities; CATI = computer-assisted telephone interviewing; ECUs=Extended Care Units; Hosp = Hospitals; NHs = Nursing Homes; RG = Registries

## Discussion

The systematic review initially identified 1,302 peer-reviewed journal articles published in the last 18 years. Detailed analysis identified only 22 empirical studies conducted in eight different countries that met the review criteria. Most articles focus on all types of abuse rather than just physical and psychological abuse. The majority of studies examine abuse from the staff perspective, with few reporting from residents, relatives and community members. Researchers have utilised study designs to include not only staff reporting abuse but other sources such as residents incorporating clinician signs of abuse, relatives and the general public. Similarly, to other reviews, we report that relatives, followed by staff tend to report the highest level of observed abuse, while resident reports the lowest abuse [12]. Measurement tools used via registries also produce low prevalence because they tend to report extreme cases of physical signs and reported abuse, however this tends to be limited to physical or sexual abuse and/or neglect.

The main aim of this review was to illustrate and critique methodologies used within the field. We identified a heterogeneity of definitions of abuse, variations of who reported abuse, a wide range of measurement tools and recall periods. There was little comparability between studies and variable study quality. The inconsistencies and poor quality make it difficult to synthesis findings, and not possible to establish the prevalence of abuse rates.

Among the 22 studies in the review, there was no consistency in presenting the study’s participants or cohort characteristics, making it difficult to conduct comparability or understanding individual study’s generalisability. The majority of cohort studies described their participants characteristics, either staff, residents or community members, using one characteristic. This point reveals there is no agreement, within or across countries, about what and how characteristics should be reported.

There was also no consistency across the 22 studies in methods and measurement tools used for investigating staff abuse among residents. Only six [36, 45, 46, 49, 50, 52] of the 22 studies had used three previously developed methodologies [35, 51] to measure older adult abuse, however, modifications were made to these original questionnaires, impacting the ability to compare findings. There were variations with recruitment methods resulting in different sample sizes and a lack of consistency in who was reporting the abuse, concluding with differences in findings. Only two studies utilised an independent researcher to personally distribute the questionnaire to staff [24] or interviewed residents face-to-face as an inpatient admitted to hospital for reasons unrelated to an incident of abuse [42] avoiding explicit bias in data analysis. Furthermore, only one study reported a prevalence of ‘self-reported’, ‘observed’, ‘committed’ or ‘experienced’ forms of older abuse by both staff and residents [47]. Study designs that focus on staff or residents reporting abuse to other staff members or facility managers, deter disclosure in their responses or create stigma and blame among staff who have witness or committed abuse, resulting in underestimated rates of abuse [46, 51]. Anonymity of those who distribute the survey, conduct interviews or examinations will reduce bias and improve reliability of the study’s findings [24, 35, 42].

It is evident that despite an increased interest in older adult abuse, as previous authors have cited, there has been minimal progress in standardising abuse measurements nationally nor globally [11]. This point highlights the unmet need to generate a robust standardized prevalence measurement tool of all types of older abuse, for use at national and global levels [6]. Instead of developing a modified questionnaire or survey, future research should focus on external validating current questionnaires.

Finally, the overall methodological assessment of the cohort of studies was poor, with only four of the 22 studies, meeting the standard expected by Boyce’s [18] prevalence study criteria. The individual studies themselves are accreditable. Heterogeneity in methodology is not valid or creditable to draw conclusions in the understanding prevalence of older adult abuse on a national nor global level. Boyce’s tool, the most generic one available, was not designed for this field and may therefore have limited the findings.

From this review, the most appropriate methodological choice for measuring older adult abuse in institutional settings would be Malmedal’s et al.(2009) [51] original 42-item questionnaire, however this is based on limited evidence, a high-quality assessment score and repeatability of the measurement tool in three studies [45, 50, 51], exhibiting a close to consistency in results. Thus, the analysis has revealed that to improve the knowledge base, there is a need for testing consistency in methods and measurement tools used for investigating staff abuse among residents. This includes greater participation from all stakeholders in research [46], and a standardised, comprehensive set of tools and data elements to be utilised. The WHO definitions provide a basis upon which these resources can be established [49]. This approach will enable accurate measurement of abuse and promote construct validity and reliability measurement tools on abuse of older adults. The proposed resources will assist in implementing effective workplace management programs to tailor associated risk factors of abuse within institutionalised care. These resources could be developed by a global consortium of experts and patient representatives, similar to internationally established methodologies in other health fields, including clinical and psychological topics [56]. Additionally, there is a need to establish a methodological quality assessment tool specific for institutionalised care to determine the level of quality of evidence. This work could take direction from that by Giannakopoulos et al. (2012) [57] and Shamliyan et al.(2010) [58] who developed instruments measuring the quality of studies examining the prevalence of disorders and diagnostic protocols or rates and risk factors for diseases. Gerontology researchers can further develop the evidence base by undertaking translational research projects. These studies will address individual organisational problems, educate, and improve staff’s understanding and identification of abuse behaviours [59, 60], and provide the broader industry policy direction [8]. All outcomes which will contribute to improvements in residents’ quality of life, safety and quality of care, and staff wellbeing – together which contribute to the quadruple aim in healthcare [61].

As a key step towards improving the evidence base and establishing standardised research tools identified above, we have developed the Aged Care Abuse Research Checklist (ACARC) (Table 3). This tool has been derived from the 22 empirical studies key strengths [19, 24, 30, 31, 35-52] and is designed to improve the methodological quality and research rigor for future studies. The ACARC comprises 11 points covering study design (2), methodology (6), results (2) and publication (1). The widespread use of ACARC can promote researchers’ engagement in collecting prevalence data on aged care abuse on national and international scales.

**Table 3:** Aged Care Abuse Research Checklist (ACARC)

| Element | Key questions |
| --- | --- |
| Study design | 1) Does the study recruit a representative cross-sectional cohort of participants using a random selection of nursing homes [35, 42] or via a professional registration list? [35] |
|  | 2) Does the study include sub representative groups within institutionalised care settings, from impaired to non-cognitive functioning residents among cognitive functioning residents? [45] |
| Methodology | 3) Does the study conduct data collection discretely with the use of an independent researcher? [24, 42] |
|  | 4) Does the study utilise a standardised validated unmodified definition and measurement tools on abuse of older adults? [56, 62] |
|  | 5) Does the study collect 'self-reported', observed, committed or experience forms of abuse from all stakeholders, including staff, residents, relatives and community members? [46] |
|  | 6) Does the study incorporate the collection of signs and symptoms of abuse by using independent professional forensic physicians, other medical staff or an independent researcher in a timely manner? [39, 40, 42, 43, 48] |
|  | 7) Does the study collect data within an agreed recall period between - ideally between three to twelve months? [63] |
|  | 8) Does the study collect the incidence of abuse:<br>a) to measure immediate signs and reports of suspected within a timely manner; and, |
|  | b) in-depth to verify incidents of abuse and to investigate the complexities attached to these abuse acts [39, 40, 43, 48] to minimise recall bias? |
| Results | 9) Does the study report on age and sex-specific estimates with confidence intervals to allow study comparisons? [18] |
|  | 10) Does the study present data on all study participants stakeholders' characteristics, as risk factors studies have shown these variables are determinates of abuse? [16] |
| Publication | 11) Does the study publish methods and results with transparency - by providing the full questionnaire and additional study results using publisher appendices to improve the study's validity and reliability? |

### Limitations

A limitation of this review was that it did not include studies examining residential special units. These environments were excluded because of their different clinical focus and unique challenge in involving residents in research. Nevertheless, the decision may have potentially excluded methodological tools measuring higher abuse rates other than indicated in this review. There is a need to conduct a specialized review and analysis for these institutionalized settings, as these groups have different needs and demands or present findings of these subgroups within articles [42, 45, 46].

## Conclusion

The review examined research methodologies used when investigating abuse within the aged care field. The review identified a heterogeneity of definitions of abuse, variation of who reported abuse, lack of agreement on measurement tools and recall periods, and variable study quality. To develop evidence-based methodology there is a need for standardised, comprehensive resources for the field. Ideally, a global consortium could be established to determine how to consistently define, accurately measure, report, analyse, and respond to abuse. The Aged Care Abuse Research Checklist (ACARC) was developed from the review as a first step towards achieving this outcome. Doing so will normalise processes within organisations and the community, allowing early interventions to change practices and reduce the risk of recurrence. These arrangements will improve resident quality of care and workplace cultures.

## Data Availability

All relevant data are within the manuscript and its Supporting Information files.

## Acknowledgments

All those involved have been the authors

---

**S1 Fig:**
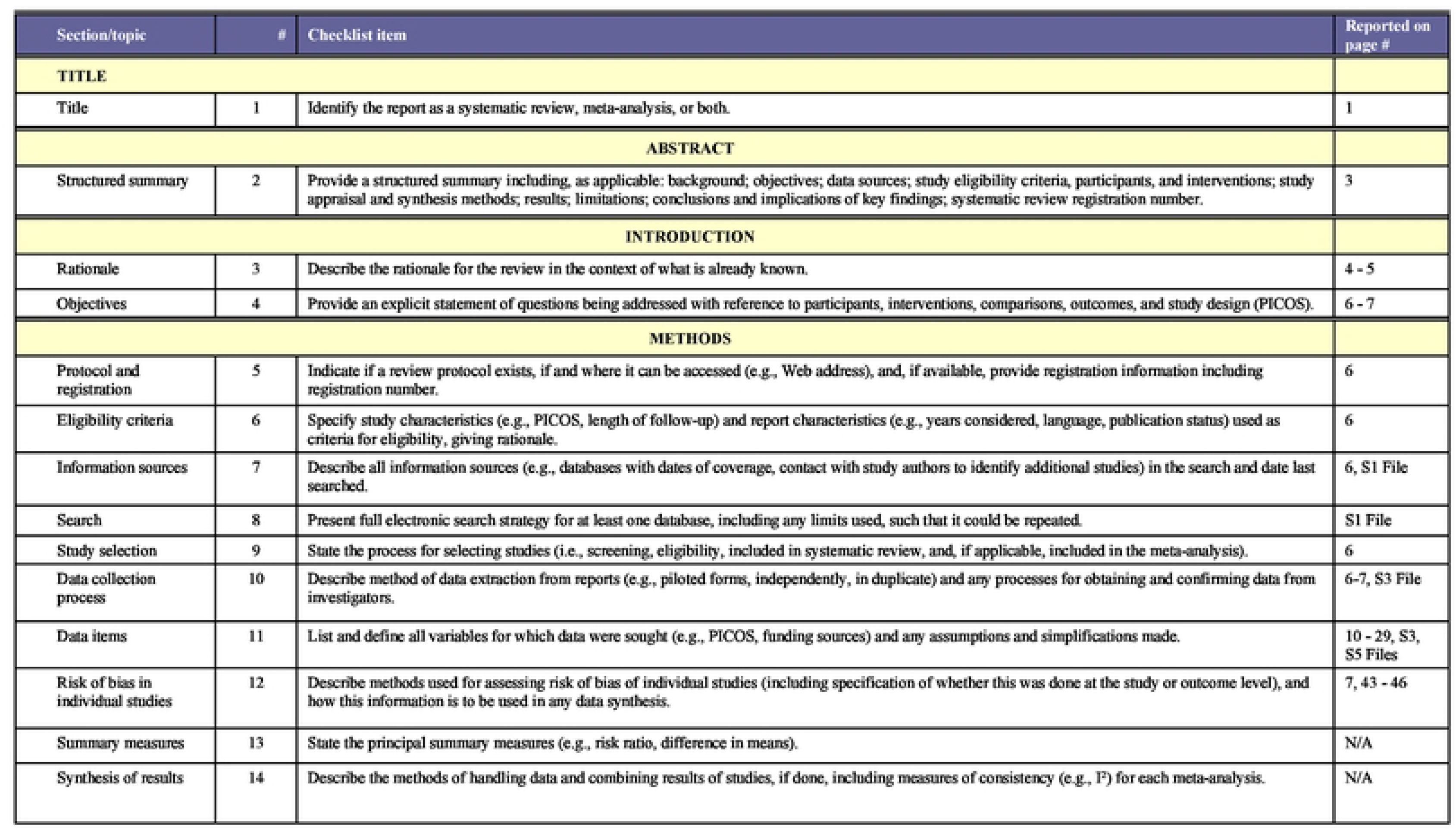
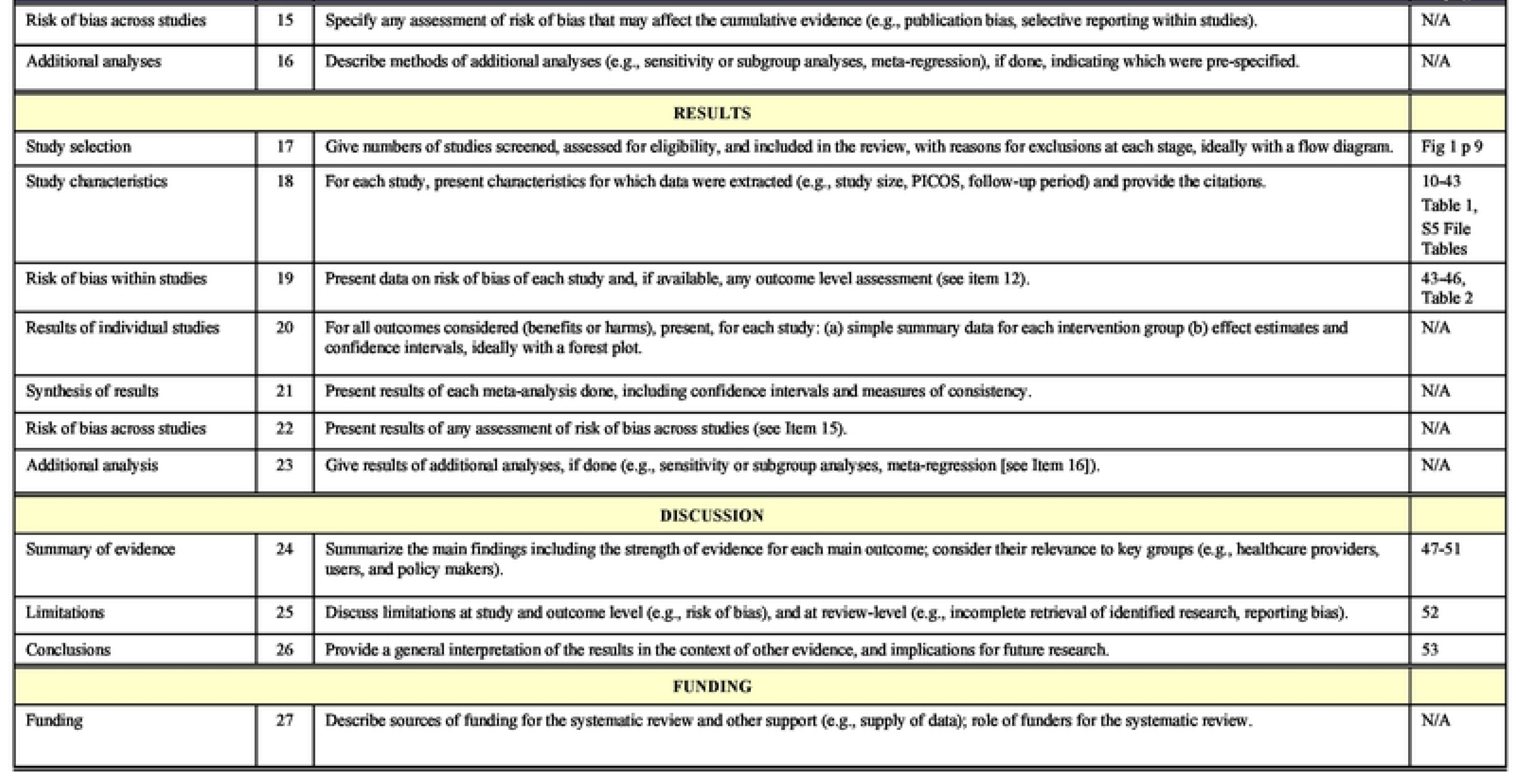
PRISMA Checklist

## Notes

Role of the Funding Source: The authors received no specific funding for this work.

Conflicts of Interest: The authors have declared that no competing interests exist.

### Competing Interest Statement

The authors have declared no competing interest.

### Clinical Protocols

https://www.crd.york.ac.uk/PROSPERO

### Funding Statement

The author(s) received no specific funding for this work.

## REFERENCES

1. Gale CR, Westbury L, Cooper C. Social isolation and loneliness as risk factors for the progression of frailty: the English Longitudinal Study of Ageing. Age Ageing. 2018;47(3):392–7. doi: 10.1093/ageing/afx188.

2. Simone L, Wettstein A, Senn O, Rosemann T, Hasler S. Types of abuse and risk factors associated with elder abuse. Swiss Med Wkly. 2016;146:w14273. doi: https://dx.doi.org/10.4414/smw.2016.14273.

3. Dyer SM, Valeri M, Arora N, Ross T, Winsall M, Tilden D, Crotty M. Review of International Systems for Long-Term Care of Older People: Report prepared for the Royal Commission into Aged Care Quality and Safety: research paper 2. [Internet] Flinders University; 2019. Available from: https://agedcare.royalcommission.gov.au/publications/Documents/research-paper-2-review-international-systems-long-term-care.pdf.

4. United Nations, Department of Economic and Social Affairs, Population Division. World Population Ageing (ST/ESA/SER.A/390). United Nations; 2015. Available from: https://www.un.org/en/development/desa/population/publications/pdf/ageing/WPA2015_Report.pdf

5. Yon Y, Ramiro-Gonzalez M, Mikton CR, Huber M, Sethi D. The prevalence of elder abuse in institutional settings: A systematic review and meta-analysis. Eur J Public Health 2019;29(1):58–67. doi: 10.1093/eurpub/cky093.

6. World Health Organization. Abuse of older people. Geneva: World Health Organization; 2022. Available from: https://www.who.int/news-room/fact-sheets/detail/abuse-of-older-people.

7. Baker JC, LeBlanc LA. Acceptability of interventions for aggressive behavior in long-term care settings: comparing ratings and hierarchical selection. Behav. Ther. 2011;42(1):30–41. doi: 10.1016/j.beth.2010.04.005.

8. Pillemer K, Burnes D, Riffin C, Lachs MS. Elder Abuse: Global Situation, Risk Factors, and Prevention Strategies. Gerontologist. 2016;56 Suppl 2(Suppl 2):S194–S205. doi: 10.1093/geront/gnw004.

9. Yon Y, Ramiro-Gonzalez M, Mikton CR, Huber M, Sethi D. The prevalence of elder abuse in institutional settings: a systematic review and meta-analysis. Eur J Public Health. 2019;29(1):58–67. doi: 10.1093/eurpub/cky093.

10. Malmedal W, Kilvik A, Steinsheim G, Botngard A. A literature review of survey instruments used to measure staff-to-resident elder abuse in residential care settings. Nurs Open. 2020;7(6):1650–60. doi: 10.1002/nop2.573.

11. De Donder L, De Witte N, Brosens D, Dierckx E, Verté D. Learning to Detect and Prevent Elder Abuse: The Need for a Valid Risk Assessment Instrument. Procedia Soc. Behav. Sci. 2015;191:1483–8. doi: 10.1016/j.sbspro.2015.04.583.

12. Hirt J, Adlbrecht L, Heinrich S, Zeller A. Staff-to-resident abuse in nursing homes: a scoping review. BMC Geriatr. 2022;22(1):563. doi: https://dx.doi.org/10.1186/s12877-022-03243-9.

13. Lindbloom EJ, Brandt J, Hough LD, Meadows SE. Elder mistreatment in the nursing home: a systematic review. JAMDA. 2007;8(9):610–6. doi: 10.1016/j.jamda.2007.09.001.

14. Page MJ, McKenzie JE, Bossuyt PM, Boutron I, Hoffmann TC, Mulrow CD, et al. The PRISMA 2020 statement: an updated guideline for reporting systematic reviews. BMJ 2021;372:n71. doi: 10.1136/bmj.n71

15. Agaliotis M, Morris T, Katz I, Greenfield D. Global Approaches to Older Abuse Research in Institutional Care Settings: A Systematic Review: PROSPERO International prospective register of systematic reviews. CRD42018055484, 18 November 2017 https://www.crd.york.ac.uk/PROSPERO.

16. Baker PR, Francis DP, Hairi NN, Othman S, Choo WY. Interventions for preventing abuse in the elderly. Cochrane Database Syst. Rev. 2016;(8):CD010321. doi: https://dx.doi.org/10.1002/14651858.CD010321.pub2.

17. Li T, Vedula S, Hadar N, Parkin C, Lau J, Dickersin K. Innovations in Data Collection, Management, and Archiving for Systematic Reviews. Ann. Intern. Med. 2015;162:287–94. doi: 10.7326/M14-1603.

18. Boyle MH. Guidelines for evaluating prevalence studies. Evid Based Ment Health. 1998;1(2):37. doi:10.1136/ebmh.1.2.37

19. Griffore RJ, Barboza GE, Mastin T, Oehmke J, Schiamberg LB, Post LA. Family members’ reports of abuse in Michigan nursing homes. J. Elder Abuse Negl. 2009;21(2):105–14. doi: 10.1080/08946560902779910.

20. Page C, Conner T, Prokhorov A, Fang Y, Post L. The effect of care setting on elder abuse: results from a Michigan survey. J. Elder Abuse Negl. 2009;21(3):239–52. doi: 10.1080/08946560902997553.

21. Post L, Page C, Conner T, Prokhorov A, Yu F, Biroscak BJ. Elder Abuse in Long-Term Care: Types, Patterns, and Risk Factors. Res Aging. 2010;32(3):323–48. doi: 10.1177/0164027509357705.

22. Schiamberg LB, Oehmke J, Zhang Z, Barboza GE, Griffore RJ, Von Heydrich L, et al. Physical abuse of older adults in nursing homes: a random sample survey of adults with an elderly family member in a nursing home. J. Elder Abuse Negl. 2012;24(1):65–83. doi: 10.1080/08946566.2011.608056.

23. Zhang Z, Schiamberg LB, Oehmke J, Barboza GE, Griffore RJ, Post LA, et al. Neglect of older adults in Michigan nursing homes. J. Elder Abuse Negl. 2011;23(1):58–74.

24. Ben Natan M, Lowenstein A, Eisikovits Z. Psycho-social factors affecting elders’ maltreatment in long-term care facilities. Int. Nurs. Rev. 2010;57(1):113–20. doi: 10.1111/j.1466-7657.2009.00771.x. PubMed PMID: 2010-02627-020.

25. Ben Natan M, Ariela L. Study of factors that affect abuse of older people in nursing homes. Nurs. Manag. (Harrow). 2010;17(8):20–4. doi: 10.7748/nm2010.12.17.8.20.c8143.

26. Moore S. Abuse of residents in nursing homes: Results of a staff questionnaire. Nurs Times. 2017;113(2):29–33.

27. Moore S. You can lead a horse to water but you can’t make it drink: How effective is staff training in the prevention of abuse of adults? J. Adult Prot. 2017;19(5):297–308. doi: 10.1108/JAP-03-2017-0008.

28. Moore S. Oops! Its happened again! Evidence of the continuing abuse of older people in care homes. J. Adult Prot. 2018;20(1):33–46. https://doi.org/10.1108/JAP-06-2017-0026.

29. Moore S. The road goes ever on: evidence of the continuing abuse of older people in care homes. Working with Older People. 2019;23(3):152–66. https://doi.org/10.1108/WWOP-06-2019-0014.

30. Moore S. Safeguarding vulnerable older people: a job for life? J. Adult Prot. 2016;18(4):214–28. https://doi.org/10.1108/WWOP-06-2019-0014..

31. Moore S. The sound of silence: Evidence of the continuing under reporting of abuse in care homes. J. Adult Prot. 2020;22(1):35–48. doi: http://dx.doi.org/10.1108/JAP-08-2019-0027.

32. Lafferty A, Drennan J, Treacy P, Lyons I, O’Loughlin A, Fealy G. Researching elder abuse in Irish residential care settings for older people: Results from a pilot study. Ir. J. Med. Sci.. 2011;180:S323. doi: http://dx.doi.org/10.1007/s11845-011-0742-0.

33. Wang JJ. Psychological abuse behavior exhibited by caregivers in the care of the elderly and correlated factors in long-term care facilities in Taiwan. J Nurs Res. 2005;13(4):271–80.

34. Neuberg M, Pudmej Esegovic V, Krizaj M, Cikac T, Mestrovic T. Abuse and neglect of older people in health facilities from the perspective of nursing professionals: A cross-sectional study from Croatia. Int. J. Older People Nurs. 2022;17(6):e12484. doi: https://dx.doi.org/10.1111/opn.12484.

35. Castle N. Nurse aides’ reports of resident abuse in nursing homes. J Appl Gerontol 2012;31(3):402–22. doi: http://dx.doi.org/10.1177/0733464810389174.

36. Castle N, Beach S. Elder abuse in assisted living. J Appl Gerontol. 2013;32(2):248–67. doi: https://doi.org/10.1177/0733464811418094

37. Friedman L, Avila S, Friedman D, Meltzer W. Association between Type of Residence and Clinical Signs of Neglect in Older Adults. Gerontol. 2019;65(1):30–9. doi: 10.1159/000492029

38. Phillips LR, Ziminski C. The public health nursing role in elder neglect in assisted living facilities. Public Health Nurs. 2012;29(6):499–509. doi: https://dx.doi.org/10.1111/j.1525-1446.2012.01029.x.

39. Teaster PB, Ramsey-Klawsnik H, Abner EL, Kim S. The Sexual Victimization of Older Women Living in Nursing Homes. J. Elder Abuse Negl. 2015;27(4-5):392–409. doi: https://dx.doi.org/10.1080/08946566.2015.1082453.

40. Teaster PB, Ramsey-Klawsnik H, Mendiondo MS, Abner E, Cecil K, Tooms M. From behind the shadows: a profile of the sexual abuse of older men residing in nursing homes. J. Elder Abuse Negl. 2007;19(1-2):29–45, table of contents. doi: https://dx.doi.org/10.1300/J084v19n01_03.

41. McCool JJ, Jogerst GJ, Daly JM, Xu Y. Multidisciplinary reports of nursing home mistreatment. JAMDA. 2009;10(3):174–80. doi: 10.1016/j.jamda.2008.09.005.

42. Cohen M, Halevy-Levin S, Gagin R, Priltuzky D, Friedman G. Elder abuse in long-term care residences and the risk indicators. Ageing Soc. 2010;30:1027–40. doi: 10.1017/s0144686x10000188.

43. Smith D, Cunningham N, Willoughby M, Young C, Odell M, Ibrahim J, et al. The epidemiology of sexual assault of older female nursing home residents, in Victoria Australia, between 2000 and 2015. Leg Med (Tokyo). 2019;36:89–95. doi: 10.1016/j.legalmed.2018.11.006.

44. Smith DE, Wright MT, Ibrahim JE. Aged care nurses’ perception of unwanted sexual behaviour in Australian residential aged care services. Australas. J. Ageing. 2022;41(1):153–9. doi: 10.1111/ajag.13014.

45. Blumenfeld Arens O, Fierz K, Zuniga F. Elder abuse in nursing homes: Do special care units make a difference? A secondary data analysis of the Swiss Nursing Homes Human Resources Project. Gerontol. 2017;63(2):169–79. doi: http://dx.doi.org/10.1159/000450787.

46. Botngård A, Eide AH, Mosqueda L, Malmedal W. Elder abuse in Norwegian nursing homes: a cross-sectional exploratory study. BMC Health Serv. Res. 2020;20(1):9. doi: https://doi.org/10.1186/s12913-019-4861-z

47. Buzgová R, Ivanová K. Violation of ethical principles in institutional care for older people. Nurs. Ethics 2011;18(1):64–78. doi: 10.1177/0969733010385529.

48. Frazão SL, Correia AM, Norton P, Magalhaes T. Physical abuse against elderly persons in institutional settings. J Forensic Leg Med. 2015;36:54–60. doi: https://dx.doi.org/10.1016/j.jflm.2015.09.002.

49. Gil AP, Capelas ML. Elder abuse and neglect in nursing homes as a reciprocal process: the view from the perspective of care workers. J. Adult Prot. 2022;24(1):22–42. doi: 10.1108/JAP-06-2021-0021.

50. Habjanič A, Lahe D. Are frail older people less exposed to abuse in nursing homes as compared to community-based settings? Statistical analysis of Slovenian data. Arch Gerontol Geriatr. 2012;54(3):e261–70. doi: https://dx.doi.org/10.1016/j.archger.2011.07.006.

51. Malmedal W, Ingebrigtsen O, Saveman B. Inadequate care in Norwegian nursing homes -- as reported by nursing staff. Scand. J. Caring Sci. 2009;23(2):231–42. doi: 10.1111/j.1471-6712.2008.00611.x

52. Neuberg M, Zeleznik D, Mestrovic T, Ribic R, Kozina G. Is the burnout syndrome associated with elder mistreatment in nursing homes: Results of a cross-sectional study among nurses. Arhiv za Higijenu Rada i Toksikologiju. 2017;68(3):190–7. doi:10.1515/aiht-2017-68-2982.

53. Drennan J, Lafferty, A., Treacy, M.P., Fealy, G., Phelan, A., Lyons, I. Hall, P. Older People in Residential Care Settings: Results of a National Survey of Staff-Resident Interactions and Conflicts. NCPOP, University College Dublin; 2012. Available from: https://www.lenus.ie/handle/10147/301725.

54. Daly JM, Jogerst GJ. Association of knowledge of adult protective services legislation with rates of reporting of abuse in Iowa nursing homes. JAMDA. 2005;6(2):113–20. doi: 10.1016/j.jamda.2005.01.005.

55. Daly JM, Jogerst GJ. Readability and Content of Elder Abuse Instruments. J. Elder Abuse Negl. 2005;17(4):31–52. doi: 10.1300/J084v17n04_03.

56. Bjelic-Radisic V, Cardoso F, Cameron D, Brain E, Kuljanic K, da Costa RA, et al. An international update of the EORTC questionnaire for assessing quality of life in breast cancer patients: EORTC QLQ-BR45. Ann. Oncol. 2020;31(2):283–8. doi: 0.1016/j.annonc.2019.10.027.

57. Giannakopoulos NN, Rammelsberg P, Eberhard L, Schmitter M. A new instrument for assessing the quality of studies on prevalence. Clin Oral Investig. 2012;16(3):781–8. doi: 10.1007/s00784-011-0557-4.

58. Shamliyan T, Kane RL, Dickinson S. A systematic review of tools used to assess the quality of observational studies that examine incidence or prevalence and risk factors for diseases. Journal of Clinical Epidemiology. 2010;63(10):1061–70. doi: https://doi.org/10.1016/j.jclinepi.2010.04.014.

59. Myhre J, Saga S, Malmedal W, Ostaszkiewicz J, Nakrem S. Elder abuse and neglect: an overlooked patient safety issue. A focus group study of nursing home leaders’ perceptions of elder abuse and neglect. BMC Health Serv. Res. 2020;20(1):199. doi: 10.1186/s12913-020-5047-4.

60. Radermacher H, Toh YL, Western D, Coles J, Goeman D, Lowthian J. Staff conceptualisations of elder abuse in residential aged care: A rapid review. Australas J Ageing. 2018;37(4):254–67. doi: 10.1111/ajag.12565.

61. World Health Organization. Continuity and coordination of care: a practice brief to support implementation of the WHO Framework on integrated people-centred health services. License: CC BY-NC-SA 3.0 IGO. [Internet]. Geneva: World Health Organization; 2018. [cited 2021 Sept 27]. Available from: https://apps.who.int/iris/handle/10665/274628.

62. Kelley TA. International Consortium for Health Outcomes Measurement (ICHOM). Trials. 2015;16(3):O4. https://doi.org/10.1186/1745-6215-16-S3-O4.

63. Beach SR, Carpenter CR, Rosen T, Sharps P, Gelles R. Screening and detection of elder abuse: Research opportunities and lessons learned from emergency geriatric care, intimate partner violence, and child abuse. J. Elder Abuse Negl. 2016;28(4-5):185–216. doi: 10.1080/08946566.2016.1229241.

